# The Bacterial and Viral Complexity of Postinfectious Hydrocephalus in Uganda

**DOI:** 10.1101/2020.08.03.20167544

**Authors:** Joseph N. Paulson, Brent L. Williams, Christine Hehnly, Nischay Mishra, Shamim A. Sinnar, Lijun Zhang, Paddy Ssentongo, Edith Mbabazi-Kabachelor, Dona S. S. Wijetunge, Benjamin von Bredow, Ronnie Mulondo, Julius Kiwanuka, Francis Bajunirwe, Joel Bazira, Lisa M. Bebell, Kathy Burgoine, Mara Couto-Rodriguez, Jessica E. Ericson, Tim Erickson, Matthew Ferrari, Melissa Gladstone, Cheng Guo, Murali Haran, Mady Hornig, Albert M. Isaacs, Brian Nsubuga Kaaya, Sheila M. Kangere, Abhaya V. Kulkarni, Elias Kumbakumba, Xiaoxiao Li, David D. Limbrick, Joshua Magombe, Sarah U. Morton, John Mugamba, James Ng, Peter Olupot-Olupot, Justin Onen, Mallory R. Peterson, Farrah Roy, Kathryn Sheldon, Reid Townsend, Andrew D. Weeks, Andrew J. Whalen, John Quackenbush, Peter Ssenyonga, Michael Y. Galperin, Mathieu Almeida, Hannah Atkins, Benjamin C. Warf, W. Ian Lipkin, James R. Broach, Steven J. Schiff

**Affiliations:** Department of Biostatistics, Product Development, Genentech Inc.; Center for Infection and Immunity, Mailman School of Public Health, Columbia University; Institute for Personalized Medicine, Dept. of Biochemistry and Molecular Biology, The Pennsylvania State University College of Medicine; Center for Neural Engineering, The Pennsylvania State University; Department of Medicine, The Pennsylvania State University College of Medicine; Department of Engineering Science and Mechanics, The Pennsylvania State University; Department of Public Health Sciences, The Pennsylvania State University College of Medicine; CURE Children’s Hospital of Uganda; Department of Pathology, The Pennsylvania State University College of Medicine; Department of Pediatrics, Mbarara University of Science and Technology; Department of Epidemiology, Mbarara University of Science and Technology; Department of Microbiology, Mbarara University of Science and Technology; MGH Medical Practice Evaluation Center, Division of Infectious Diseases, and Center for Global Health; Neonatal Unit, Department of Paediatrics and Child Health, Mbale Regional Referral Hospital, Mbale, Uganda; Mbale Clinical Research Institute, Mbale Regional Referral Hospital, Mbale, Uganda; University of Liverpool, Liverpool, UK; Biotia, 100 6th avenue, New York, NY, USA; Division of Pediatric Infectious Disease, The Pennsylvania State University College of Medicine; The Center for Infectious Disease Dynamics, The Pennsylvania State University; Departments of Biology and Statistics, The Pennsylvania State University; Institute for Translational Medicine, University of Liverpool; Department of Statistics, The Pennsylvania State University; Department of Epidemiology, Columbia University Mailman School of Public Health; Department of Neuroscience, Washington University School of Medicine; Department of Neurosurgery, University of Toronto; Department of Neurological Surgery, Washington University School of Medicine; Division of Newborn Medicine, Boston Children’s Hospital and Department of Pediatrics, Harvard Medical School; Busitema University, Faculty of Health Sciences, Mbale Campus 2; Department of Biostatistics, Harvard T.H. Chan School of Public Health; Department of Medicine, Washington University School of Medicine; Department of Women’s and Children’s Health, University of Liverpool and Liverpool Women’s Hospital for Liverpool Health Partners, Liverpool, UK; Department of Mechanical Engineering, The Pennsylvania State University; National Center for Biotechnology Information, National Library of Medicine, National Institutes of Health; MGP MetaGénoPolis, INRA, Université Paris-Saclay; Department of Comparative Medicine, The Pennsylvania State University College of Medicine; Dept. of Neurosurgery, Boston Children’s Hospital, Harvard Medical School; Department of Neurosurgery, The Pennsylvania State University College of Medicine; Department of Physics, The Pennsylvania State University

## Abstract

Postinfectious hydrocephalus (PIH), often following neonatal sepsis, is the most common cause of pediatric hydrocephalus world-wide, yet the microbial pathogens remain uncharacterized. Characterization of the microbial agents causing PIH would lead to an emphasis shift from surgical palliation of cerebrospinal fluid (CSF) accumulation to prevention. We examined blood and CSF from 100 consecutive cases of PIH and control cases of non-postinfectious hydrocephalus (NPIH) in infants in Uganda. Genomic testing was undertaken for bacterial, fungal, and parasitic DNA, DNA and RNA sequencing for viral identification, and extensive bacterial culture recovery. We uncovered a major contribution to PIH from *Paenibacillus*, upon a background of frequent cytomegalovirus (CMV) infection. CMV was only found in CSF in PIH cases. A facultatively anaerobic isolate was recovered. Assembly of the genome revealed a strain of *P. thiaminolyticus*. In mice, this isolate designated strain *Mbale*, was lethal in contrast with the benign reference strain. These findings point to the value of an unbiased pan-microbial approach to characterize PIH in settings where the organisms remain unknown, and enables a pathway towards more optimal treatment and prevention of the proximate neonatal infections.

**One Sentence Summary:** We have discovered a novel strain of bacteria upon a frequent viral background underlying postinfectious hydrocephalus in Uganda.

## Introduction

Hydrocephalus is the most common indication for neurosurgery in children. Of the estimated 400,000 new cases each year, about half are estimated to be postinfectious, with the largest number of cases in low- and middle-income countries, especially sub-Saharan Africa (*1*). Neonatal sepsis (*2*) often precedes postinfectious hydrocephalus (PIH) (*3*), although the manifestations of hydrocephalus typically emerge in the months following the neonatal period as sufficient cerebrospinal fluid (CSF) accumulates so that cranial expansion garners medical attention. Thus, although these infants will typically die in early childhood without advanced surgical management, they are omitted from neonatal mortality surveillance (*4*).

The spectrum of microbial agents that underlie PIH remains poorly characterized. It is known that seasonal *Neisseria* epidemics can produce such cases within the African meningitis belt (*5*), and there have been reports of a tendency towards gram negative coliform bacteria in infants in other Southern (*6*) and Eastern (*7*) African locations where *Neisseria* is uncommon.Nevertheless, there has never been a well-controlled examination of the agents underlying PIH, and no knowledge of the roles that viruses, parasites, or fungi might play in addition to bacteria. If the microbial agents causing PIH were better characterized, emphasis could shift from high technology palliation of CSF accumulation (*8*) to prevention.

In this study, we examined blood and CSF from 100 consecutive cases of PIH and control non-postinfectious hydrocephalus (NPIH) in infants under 3 months of age at the CURE Children’s Hospital of Uganda (CCHU) in Mbale, Uganda. Since 2001, this pediatric neurosurgical hospital has treated thousands of PIH and NPIH cases, with nearly uniformly negative recovery of putative pathogens through standard bacterial culture. In this study, we gathered high quality blood and CSF samples for molecular analysis, and comprehensive testing was undertaken for bacterial, fungal, and parasitic DNA, genomic and RNA transcript sequencing for viruses, in addition to extensive bacterial culture recovery efforts for taxonomic identification, genome assembly and virulence characterization.

## Results

### Demographics & clinical characteristics

Between March and November 2016, 115 consecutive patients were screened, and 15 were excluded for the following reasons: 8 did not give consent, 6 lived outside of Uganda (South Sudan and Kenya), and 1 weighed less than 2.5 kg. A total of 100 patients 3 months of age or younger with hydrocephalus were enrolled; 64 with PIH and 36 with NPIH. PIH patients were on average several weeks older than NPIH patients (whose disorders were generally recognized at birth), and had higher peripheral and CSF white blood cell counts, lower blood hemoglobin and hematocrit levels, but were evenly distributed by sex and HIV exposure status (Table 1).

**Table 1:** Demographics and clinical characteristics of NPIH and PIH cohort. Patient demographics and Clinical Characteristics. Comparison of demographic and clinical attributes between two groups: Postinfectious hydrocephalus (PIH, n=64) and non-postinfectious hydrocephalus (NPIH, n=36). PIH patients were older (66 days vs 43 days, p<0.0001), had higher peripheral and cerebrospinal fluid (CSF) white blood cell (WBC) counts, and were more likely to be anemic (hemoglobin 10.7g/dL vs 13.0 g/dL, p<0.0001) compared with NPIH patients. Preoperative computed tomography (CT) scans were available for 98 subjects. Of these, the PIH group was more likely to have a higher CT scan score reflective of brain abscess, calcifications, loculations and septations (p<0.0001). There were no significant differences in gender and human immunodeficiency virus (HIV) exposure frequencies between the groups. Continuous demographic variables were evaluated using the non-parametric Wilcoxon rank-sum (2-group comparisons) and Kruskal-Wallis (>2 groups) tests following Shapiro-Wilk’s test for normality unless otherwise stated. Fisher’s exact test was performed for categorical variables.

| Characteristics | All patients<br>n=100 | PIH<br>n=64 | NPIH<br>n=36 |
| --- | --- | --- | --- |
| Age in days, mean (SD) | 57 (24) | 66 (17) | 43 (27) |
| Sex |  |  |  |
| Male (%) | 51 (51) | 35 (55) | 16 (44) |
| Female (%) | 49 (49) | 29 (45) | 20 (56) |
| Peripheral blood WBC<br>[1.0 x 10<sup>3</sup>]/μL, mean (SD) | 10.3 (3.6) | 11.0 (3.8) | 8.8 (2.6) |
| CSF WBC/μL, mean (SD)‡ | 30 (62) | 45 (74) | 5 (0.5) |
| Hemoglobin g/dl, mean (SD) | 11.5 (2.2) | 10.7 (1.3) | 13.0 (2.8) |
| Hematocrit %, mean (SD) | 36.8 (7.4) | 34.1 (4.1) | 41.6 (9.3) |
| CT scan scoring*<br>[no. and % in each category] |  |  |  |
| 0 | 32 (33) | 8 (25) | 24 (69) |
| 1 | 15 (15) | 7 (11) | 8 (23) |
| 2 | 13 (13) | 10 (16) | 3 (8) |
| 3 | 11 (11) | 11 (17) | 0 (0) |
| 4 | 27 (28) | 27 (43) | 0 (0) |
| HIV Exposure status [no. and %] |  |  |  |
| Yes | 5 (5) | 3 (5) | 2 (6) |
| No | 95 (95) | 61 (95) | 34 (94) |

\* n=98
‡ n=99

Prior to admission for hydrocephalus, 15 infants received antibiotics, but we only have records detailing antibiotic type for 2 of these 15 patients (gentamicin with ampicillin in one case and gentamicin with ceftriaxone in the other case). Three patients received antibiotics for treatment of active infection at CCHU after admission (ceftriaxone with gentamicin in one case, and ceftriaxone alone in the others). Such antibiotic treatment is adjunctive to abscess management (drainage and irrigation of larger accessible abscesses), prior to more definitive surgical treatment of hydrocephalus with endoscopic third ventriculostomy or insertion of a ventriculoperitoneal shunt (*8*).

Preoperative CT scans were scored, demonstrating that PIH patients were more likely to have evidence of CSF fluid loculations, debris within fluid spaces, ectopic calcification, and brain abscesses (Table 1 and Fig. S1). The homes of PIH patients were concentrated within central and eastern Uganda, in a swampy plateau north of Lake Victoria, and south and north of the banks of Lake Kyoga, while NPIH patients were more uniformly distributed geographically (Fig. 1 and Fig. S2, p=0.03 by linear discrimination).

**Figure 1.**
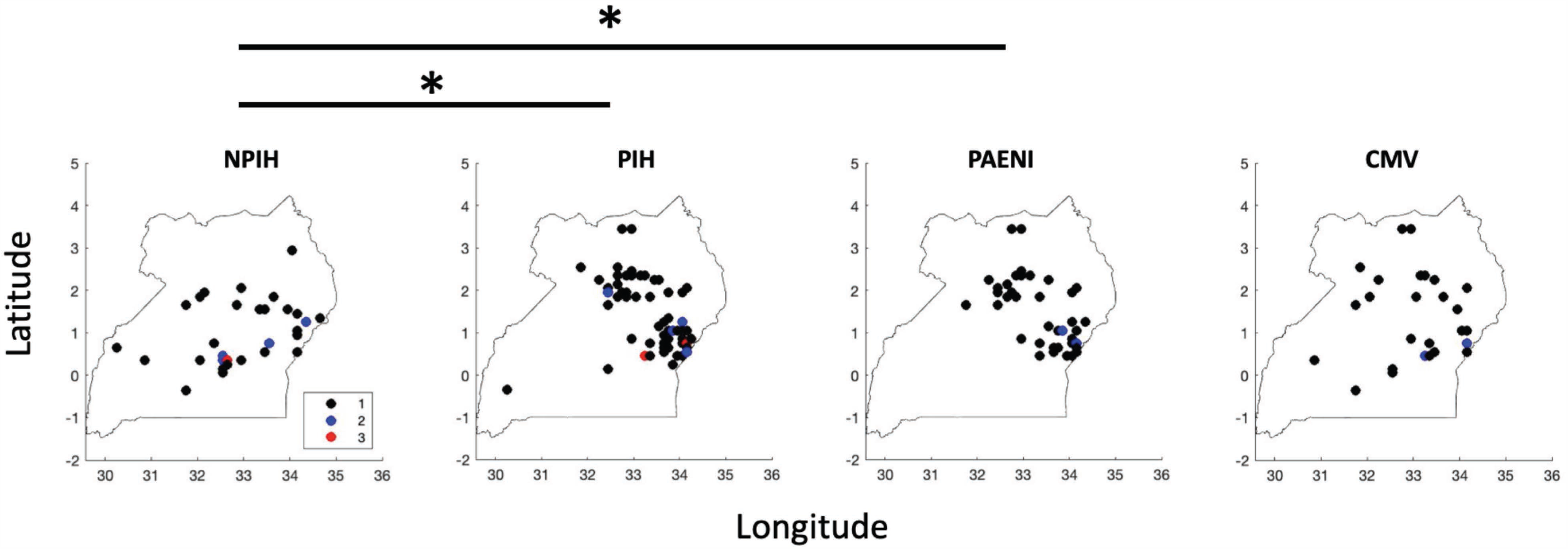
Comparative location of cases within map of Uganda. The village centroid GPS locations are mapped to the 0.1 × 0.1 degree grid frequently used in satellite rainfall estimation (*54*), and where village name was uncertain, the centroid of the administrative parish or sub-county was used. The groups shown are mapped by clinical status (NPIH and PIH), and by organism type (*Paenibacillus*, PAENI; cytomegalovirus, CMV). Both the PAENI and CMV mappings included all NPIH and PIH cases with such diagnoses. Using Fisher’s linear discrimination analysis, group comparisons by latitude and longitude mapping could significantly discriminate NPIH from PIH, or NPIH from PAENI, at the p < 0.01 level parametrically (Wilk’s lambda), and at the p= 0.03 and p=0.01 level respectively using a bootstrap method (see Figure S2). The number of cases mapped to a 0.1 × 0.1 degree grid (11 km per edge at the equator) are indicated with colored circles as 1, 2, or 3.

### Bacterial pathogen detection

For bacterial pathogen discovery, both Sanger sequencing of 16S rDNA V1-V4 region and next generation sequencing of V1-V2 and V4 regions were performed on fresh frozen and preserved specimens in two different laboratories, which enabled us to account for known variation in microbial community amplicon sequencing (*9*), and demonstrate reproducibility of our findings.

Using DNA from fresh frozen CSF, conventional PCR targeting the V1-V4 16S rDNA gene region (Table S1) revealed 16S amplification in 27/64 PIH and only 3/36 NPIH patients. V1-V4 Sanger sequencing of subcloned amplicons identified *Paenibacillus* as a predominant organism within the PIH cohort (23/64), but not within the NPIH cohort (0/36). The full results from this method are summarized in Table S2. For quantification of *Paenibacillus* in CSF, *Paenibacillus* genus-specific qPCR was performed confirming and quantifying 22 of the 23 samples that were positive for *Paenibacillus* by Sanger sequencing and identifying 4 additional positive cases (Fig. 2A and 2B). Next generation sequencing on 16S rDNA V4 region was performed on all samples from which amplification libraries could be obtained with composite MiSeq primers (26/64 PIH and 3/36 NPIH) (Fig. 2C). Only a few nucleotides distinguished *P. thiaminolyticus* and *P. popilliae* within the V1-V4 region, hindering species-level discrimination between these taxa. From the V1-V4 sequencing data a phylogenetic tree was constructed, revealing that the majority of the *Paenibacillus* V1-V4 sequences were most closely related to *P. thiaminolyticus* and *P. popilliae*, and sequences from one subject that most closely matched *P. alvei* (Fig. S3).

**Figure 2.**
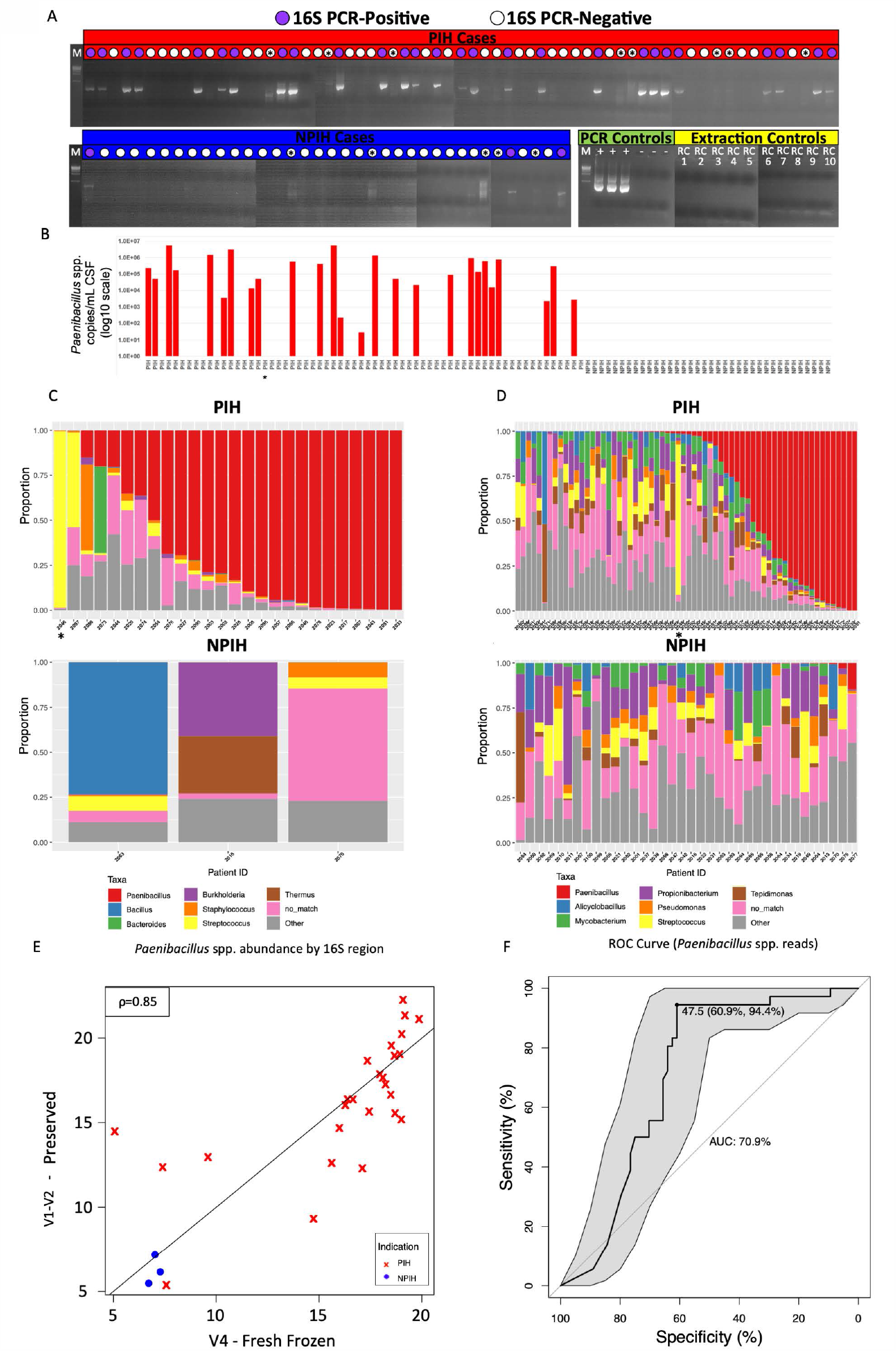
Detection and typing of bacteria using 16S rDNA. A) Agarose gels showing 16S rDNA amplification products for PIH (red) and NPIH (blue), along with PCR positive (+) and negative (-) controls (green) and 10 separate extraction reagent controls (yellow). Brightness and contrast were adjusted for gels to maximize visibility of faint bands and smears. Asterisk (*) denotes lanes where faint non-specific amplification (smears) or bands of unexpected size were observed. All amplification products including non-specific amplifications were subjected to subcloning and sequencing. B) *Paenibacillus* qPCR quantification. C-D) Stacked bar relative abundance plots of (C) 16S V4 and (D) V1-V2 regions in microbial communities for the most dominant bacteria observed within (C) 16S positive samples from (B) and in (D) all 100 samples. Star underneath PIH sample (C, D) highlights individual PIH sample with group B streptococcal infection. E) Scatterplot of cumulative sum scaling normalized *Paenibacillus* abundance for (abscissa) V4 16S sequencing of fresh frozen CSF samples and (ordinate) V1-V2 16S sequencing of biological replicates from preserved CSF samples. F) Receiver-operating-characteristic (ROC) curve using the number of *Paenibacillus* reads as the predictor for PIH or NPIH status. Area under the curve was 70.88% (95% DeLong CI = 60.61%-81.15%). Sensitivity and specificity are maximized at 47.5 reads in a given sample, consistent with the threshold employed of 50 reads.

Using DNA from samples in genomic preservative, next generation sequencing was performed on V1-V2. Overall, representative sequences from the 1,767 operational taxonomic units (OTUs) matched 159 genera. The majority of OTUs were sparsely represented except for a number of known skin flora, e.g., *Propionibacterium* spp. (Fig. 2D). Over half of the reads in 20% of the patients were attributed to the genus *Paenibacillus. Paenibacillus* spp. were present, defined as a minimum of 50 reads, in 38 PIH patients and 2 NPIH controls (Fig. 2D).

To associate taxa with infection we aggregated annotated OTUs at the genus level and performed differential abundance analysis. In performing linear regression analysis (Supplemental Methods: Differential Abundance Analysis), *Paenibacillus* was the only genus associated with PIH following multiple testing correction (Table S3). *Paenibacillus* spp. 16S rDNA abundance was used as a biomarker for classifying PIH patients and was consistent between V1-V2 and V4 (Fig. 2E). A receiver operating characteristic analysis yielded an area under the curve of 70.9% (95% DeLong CI = 60.6%-81.1%) for V1-V2 (Fig. 2F, Fig. S4A), with an optimal threshold just below 50 reads.

The spatial distribution of PIH and PIH *Paenibacillus* positive cases was significantly different from control NPIH cases (Fig. 1, Fig. S2).

Other putative pathogens detected by 16S in individual patients at high abundance included sequences consistent with *Bacillus subtilis* and *Streptococcus agalactiae* (Fig. 2C and 2D, Table S2). Diversity decreased as *Paenibacillus* abundance increased (Fig. S4B). The majority of CSF samples had similar microbial background leading to no clear visual separation of PIH and NPIH when diversity (beta) was visualized with principal coordinates analysis (PCoA) plots to reduce the dimension (Fig. S4C). Limiting to the set of OTUs annotated as *Paenibacillus* spp. revealed two or three clusters of patients with similar *Paenibacillus* abundance distributions in positive patients (Fig. S5). Further analysis on the microbial communities, including comparison of 16S regions and the characterization of isolates’ taxonomy, is described in Supplementary Methods (microbial characterization: taxonomic assignment and overview and differential abundance analysis).

### Viral pathogen detection

Utilizing the targeted viral detection capture technique VirCapSeq-VERT (*10*), we observed evidence of 11 viral strains distributed across 36% of samples — 32.8% (PIH) and 41.6% (NPIH) (Table S4). Only Human Herpesvirus 5 (cytomegalovirus, CMV) was present at substantial abundance, confirmed by requiring positive findings on at least two replicates in two different qPCR methods (Supplementary Materials, Table S4) applied to all 100 preserved CSF and blood samples. CMV was confirmed in 27/100 patients (27/99 blood: 18/64 PIH, 9/35 NPIH), but CMV was found only in the CSF in blood CMV positive PIH cases (8/100 CSF: 8/64 PIH, 0/36 NPIH) (Fig. 3A). RNA sequence data confirmed 4 cases of CMV by sequence matches to multiple mRNA transcripts (Fig. 3A).

**Figure 3.**
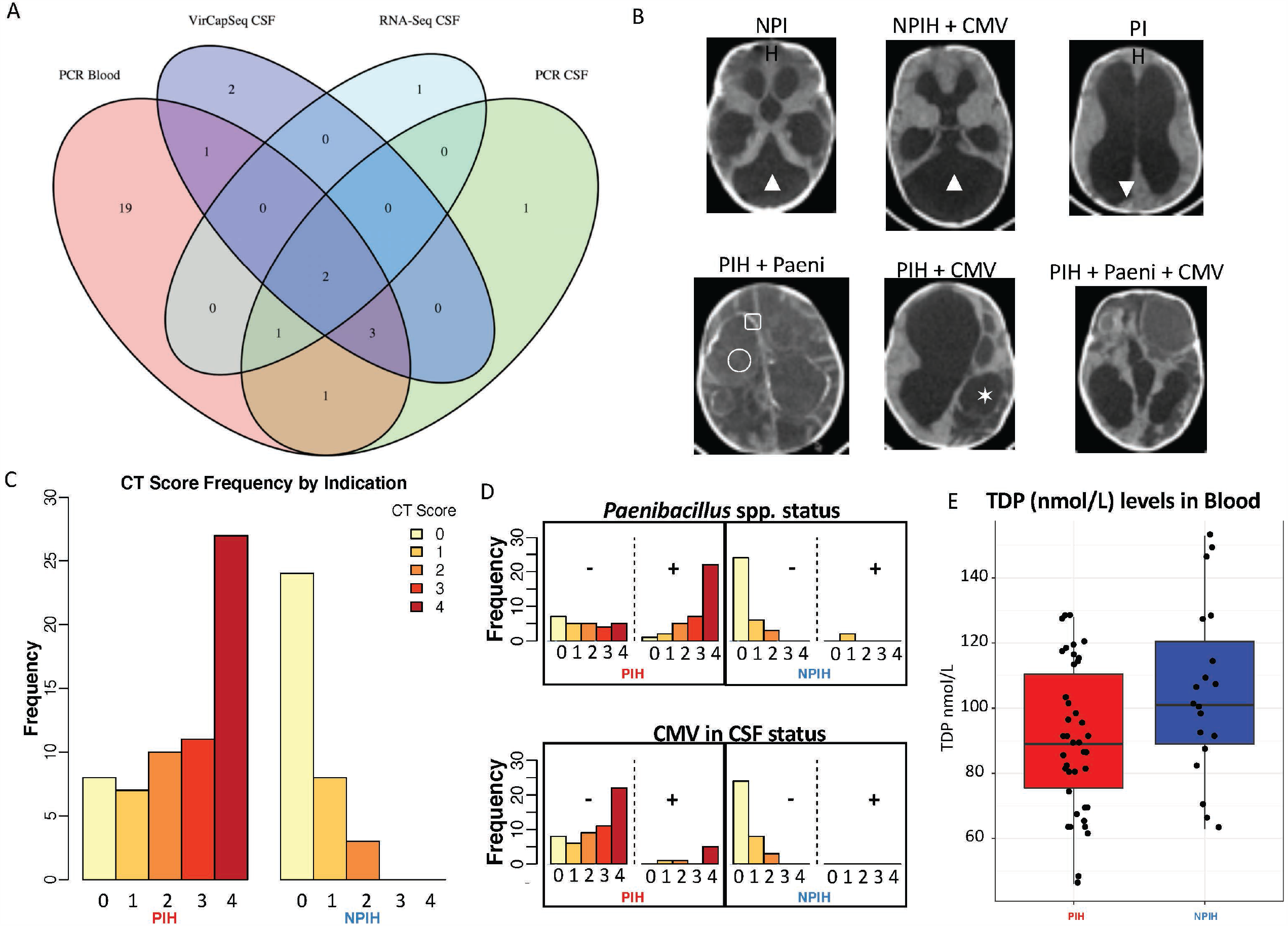
Viral detection, CT findings, thiamine levels, and infection status. A) Venn diagram of CMV detection using PCR on blood, and VirCap-Seq, RNA-Sequencing and PCR on CSF samples. B) Representative images of patients with NPIH or PIH, and CMV or *Paenibacillus* (Paeni) status. Both NPIH demonstrate Dandy-Walker cyst malformations (▲) without history or surgical findings reflective of prior infection. Note the presence in the *Paenibacillus* PIH cases of multiple loculations of CSF (✶), higher density fluid collections reflective of debris or blood within the CSF (▼), ectopic calcification within the brain (▢), and abscess formation (◯). C) CT scores stratified by clinical indication, PIH or NPIH, and D) as a function of CMV and *Paenibacillus* infection status, positive or negative respectively (see also Fig S6). E) Levels, in nmol/L, of thiamine diphosphate (TDP) in PIH (n=42) vs NPIH (n=19) cases. We observed that TDP levels are lower in PIH patients, (t-test, p<0.05). Boxplots display the median and upper and lower quartiles with whiskers forming the 1.5x the interquartile range.

The spatial distribution of CMV positive cases was not significantly different from control NPIH cases (Fig. 1, Fig. S2).

### Paenibacillus correlates with clinical signs

Several clinical measurements were positively associated with *Paenibacillus* presence including CSF cell count and CT scan scores.

In 12 months of follow-up, there were 5 deaths: 3 PIH and 2 NPIH patients. Each of the PIH deaths were in patients with *Paenibacillus*.

*Paenibacillus* spp. abundance was inversely correlated with patient age, consistent with residua from neonatal infection (Fig. 4A, Fig. S6, Kruskal-Wallis, P<0.05). Only PIH patients had high CSF cell counts (>5/μL); all were positive for *Paenibacillus* in the top abundance quartiles (Fig. 4B, Fig S6). Infants with hydrocephalus may have considerable extra weight in their heads from CSF relative to their body mass, and after calculating and subtracting excess fluid volume-for-age (*11, 12*), we found no difference between this corrected weight-for-age with hydrocephalus or *Paenibacillus* status (Fig. 4C). Seizures were more commonly reported in patients prior to admission (25 vs 13) and during hospital admission (9 vs 1) in patients who were *Paenibacillus* positive. The mean (SD) number of estimated days from initial febrile episode to when the head was noted to be growing was 21.4 (16.4) versus 29.3 (25.4) for *Paenibacillus* positive versus negative cases. Bloody CSF was noted in 15 CSF samples, but did not account for Paenibacillus positivity (1/7 positive in PIH), or the presence of CMV in CSF (0/15 positive in PIH or NPIH).

**Figure 4.**
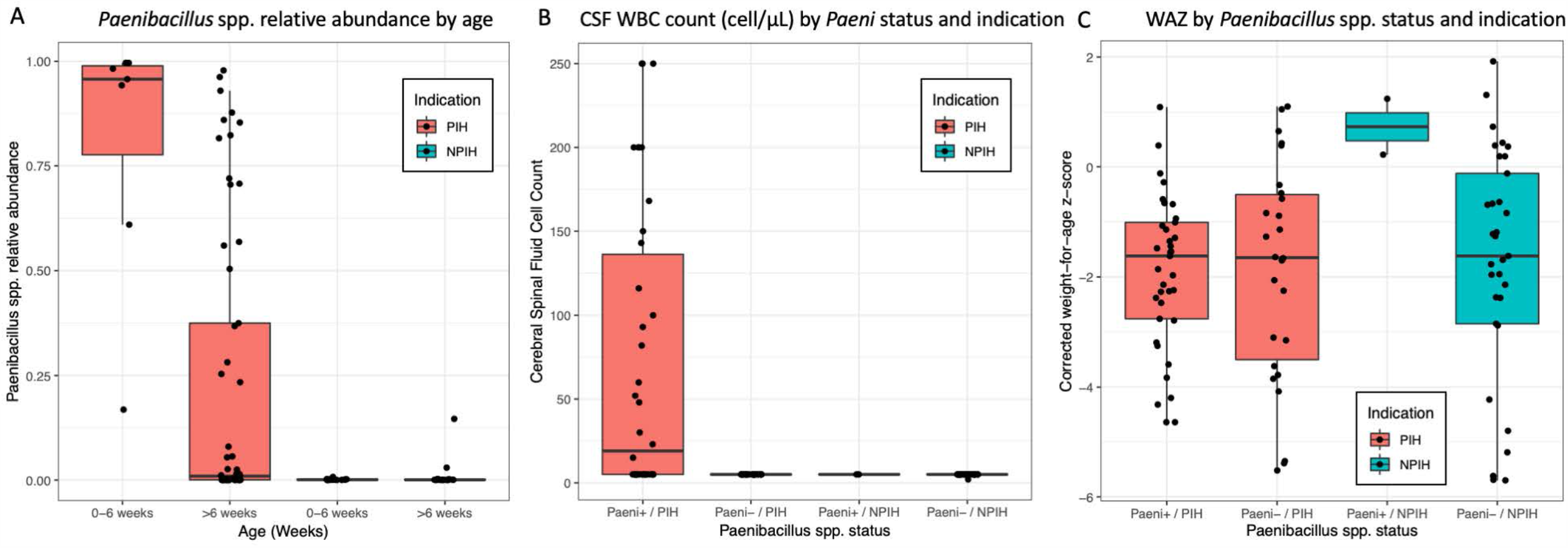
Clinical signs associate with *Paenibacillus* detection. A) Boxplots of (ordinate) log2 normalized *Paenibacillus* 16S rDNA abundance by age (categorized into 0-6 weeks and >6 weeks) and by indication (see also Fig S6). B) Boxplots of (ordinate) CSF WBC count (cell/µL) by *Paenibacillus* status (+/-) and by indication (see also Fig S6). Cell count values less than 5 were mapped to 5. Cell count values greater than 250 were mapped to 250. C) Boxplots of (ordinate) corrected weight-for-age z-scores (WAZ), after calculating and subtracting excess CSF volume-for-age, by *Paenibacillus* status (+/-) and by indication. Boxplots display the median and upper and lower quartiles with whiskers forming the 1.5x the interquartile range.

Brain image CT scan scoring representative of brain abscesses, calcifications, loculations, and debris was calculated using preoperative imaging. PIH patients without measurable *Paenibacillus* had higher scores than NPIH patients, and PIH patients positive for *Paenibacillus* had significantly higher scores compared to PIH patients without *Paenibacillus* detected (Fig. 3B, 3C, Table S5, Figures S1, S6 and S7). All of the CSF positive CMV patients were PIH patients (Table S6), and each had at least one of the four signs comprising the CT score: 6/7 fluid loculations, debris within fluid spaces, or ectopic calcification, and 5/7 abscess (Table S7). We fit an ordinal logistic regression model that included PIH vs NPIH status and *Paenibacillus* presence. Patients with PIH had increased proportional odds for high CT score, OR (95% CI) = 11.66 (4.29, 33.94) as well as *Paenibacillus* presence, OR (95% CI) = 7.6 (3.06, 19.88). Testing for CMV presence did not show increased proportional odds of high CT scan scores (Fig. 3D), OR (95% CI) = 3.30 (0.52, 29.66), when controlling for hydrocephalus etiology and *Paenibacillus* presence.

### Growth and characterization of Paenibacillus strain

From 600 initial cultures from fresh frozen CSF (Table S8), 12 isolates were recovered from 7 patients (Table S9). Two isolates of *Paenibacillus* were recovered from cultures using small volumes (50 µl) of fresh frozen patient samples, and 1 from the blind culture of a lytic anaerobic bottle (BD BACTEC). These were identified as *Paenibacillus* spp. using MALDI-TOF (Table S9). Of the three inoculum recoveries, two grew from subculture or blind culture onto solid media, identifying them as facultative anaerobes, while the third was never successfully subcultured. Marker gene analysis of the three *Paenibacillus* isolates were identified as *P. thiaminolyticus, P. amyloliticus*, and *Paenibacillus* sp. (Fig. 5, Supplementary Methods: phylogenetic tree placement). The 16S rRNA genes from the whole genome sequences of all 3 isolates were compared to the 16S amplicon V1-V2 OTU cluster representative sequences. The *P. thiaminolyticus* isolate was most similar to that of OTU 99373, the most dominant OTU annotated as *Paenibacillus* within *Paenibacillus*-positive patients (Fig. S5). This identified the *P. thiaminolyticus* strain (hereafter, strain *Mbale*) as our isolate of interest for the subsequent tests for virulence in mice. Further, we compared this strain by 16S rRNA gene similarity, average nucleotide identity (ANI) (http://enve-omics.ce.gatech.edu/ani/) (*13*), and biochemical testing against the *P. thiaminolyticus* type strain NRRL B-4156^T^ (=JCM 8360^T^,GenBank accession CP041405). The 16S rRNA genes of this isolate had 99.2%-99.4% identity, and the whole-genome average nucleotide identity (gANI) value was 97.06%, well above the 94-96% species threshold (*14*). Biochemical testing (https://apiweb.biomerieux.com, Table S10) had a 99.5-99.9% identity confirming that this isolate belongs to *P. thiaminolyticus* (*15, 16*). Antibiotic sensitivity was tested and the *Mbale* strain was broadly sensitive to common antibiotics (Table S11).

**Figure 5.**
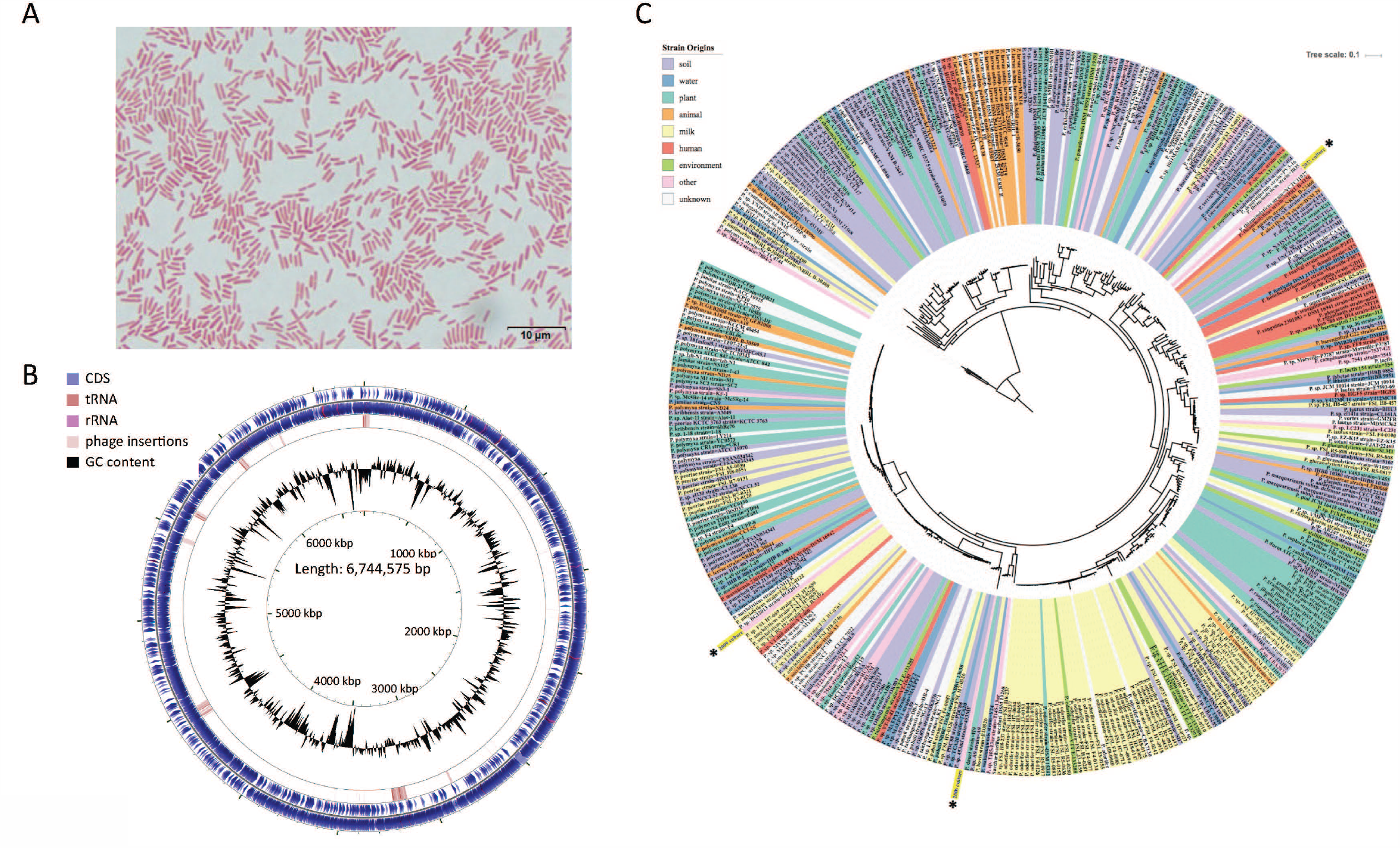
Whole genome sequencing and assembly of *P. thiaminolyticus* CSF isolate strain Mbale. From the cultured CSF samples, three isolates were identified as *Paenibacillus* (marked with asterisks in panel C). One isolate showed high 16S rDNA sequence identity to our V1-V2 and V4 sequencing results thus making it the isolate of high interest. A) *P. thiaminolyticus* Mbale Gram stain from the chocolate agar subculture of the lytic anaerobic bottle at 1,000x magnification. Weak or negative Gram staining, despite a Gram positive cell structure, is characteristic of *Paenibacillus* species (*55*). B) To classify the *P. thiaminolyticus* clinical isolate, an extensive genome analysis was performed using both long-read and next generation sequencing along with optical mapping (GenBank Accession CP041404). The resulting draft circular genome shown was created using CGView which features coding sequences (CDS), tRNA, rRNA, phage insertions and GC content (53%). C) Phylogenetic tree of *Paenibacillus* spp. based on 40 marker genes. Cultured CSF isolates are indicated by yellow marks; the isolate 2033 was renamed *P. thiaminolyticus* strain Mbale.

### Thiamine testing

We tested for thiamine deficiency assaying for thiamine diphosphate (TDP) levels in whole blood. PIH cases (positive and negative for *Paenibacillus*) had lower TDP blood levels than NPIH cases (Fig. 3E, t-test, p<0.05).

### Pathogenicity of Paenibacillus strain Mbale

Comparative virulence was assessed between the reference strain NRRL B-4156 and the strain *Mbale* in age-matched C57BL/6J mouse littermates of both sexes inoculated intraperitoneally at postnatal days 21-28. The reference strain demonstrated no adverse effects on the mice, while the strain *Mbale* produced illness in all mice (16/16), with mortality or moribund states in 15/16 (93%) of animals inoculated at a comparable concentration of colony forming units (Fig. 6, Table S12). The *Mbale* strain produced acute tubular necrosis in the kidneys, bone marrow myeloid hyperplasia, and moderate lymphocyte apoptosis in the splenic periarteriolar sheaths, but there were no significant brain lesions (Fig. 6).

**Figure 6.**
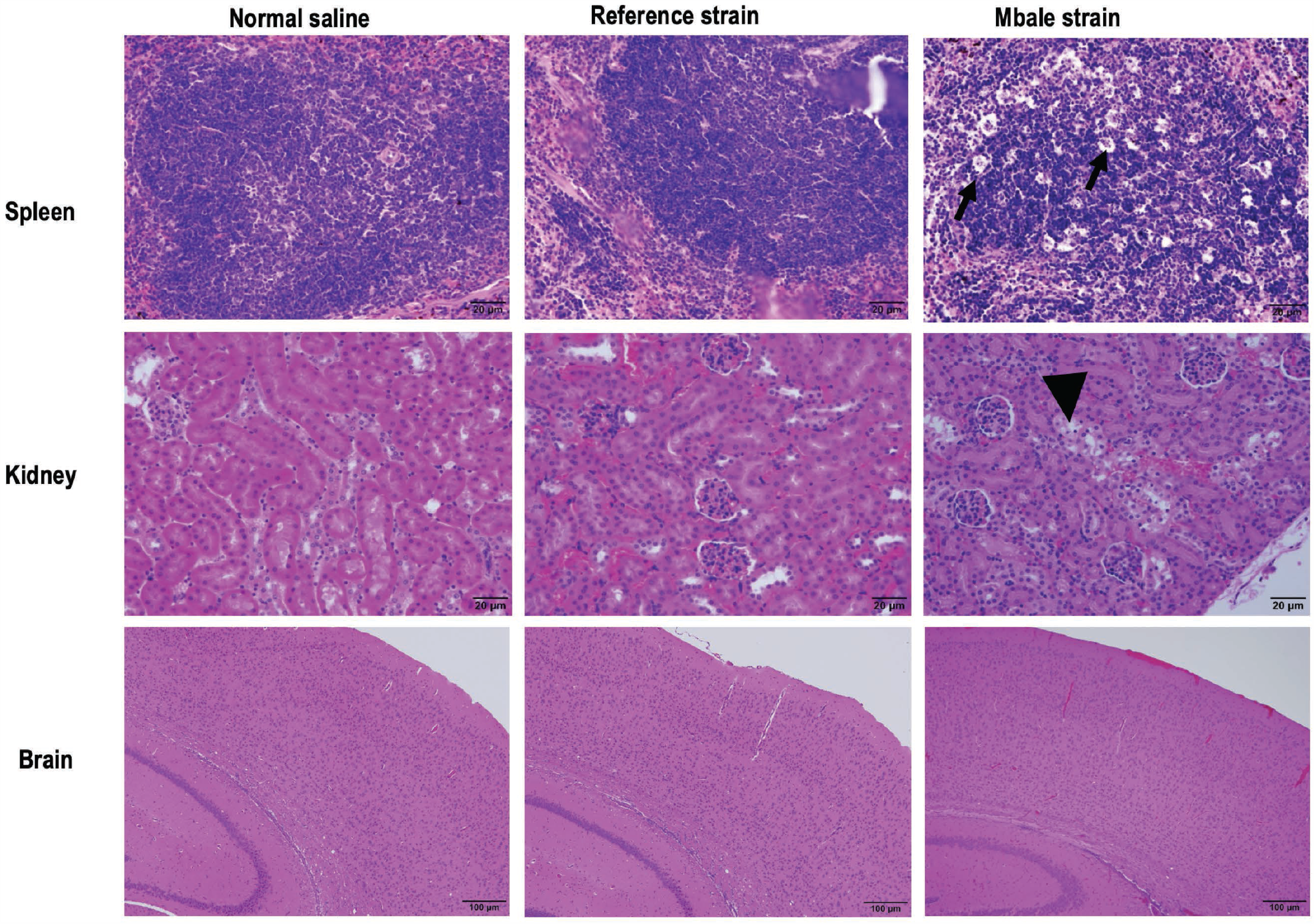
Comparative murine histology of reference and Mbale strains of *Paenibacillus thiaminolyticus*. Murine model contrasting the virulence of the *Mbale* strain of *P. thiaminolyticus* vs the reference *P. thiaminolyticus* type strain NRRL B-4156^T^ following intraperitoneal injection into C56BL/6J littermates, in comparison with saline control injections. No significant lesions are seen (top row) in the spleen of mice inoculated with normal saline (n=6) or reference strain (n=10). However, tingible body macrophage necrosis (black arrows) with intracytoplasmic apoptotic bodies were present in spleen of mice inoculated with the *Mbale* strain (9/10), suggestive of toxin induced apoptosis and a marked immune response with a high lymphocyte turnover. Likewise, the kidneys (middle row) of mice inoculated with normal saline or the reference strain had no significant lesions. In contrast, pyknotic nuclei (arrow head) of proximal tubule epithelial cells with cell sloughing and loss were observed within mice inoculated with the *Mbale* strain (9/10), indicative of acute tubular necrosis. In the brain (bottom row), there was no substantial evidence of infection-associated pathology (normal saline n=6/6, reference strain n=10/10, *Mbale* strain n=10/10).

## Discussion

Postinfectious hydrocephalus may be the largest single cause of childhood hydrocephalus and the need for neurological surgery in children worldwide. These cases are concentrated in low- and middle-income countries (*1*), and the dominant predisposing event is often neonatal sepsis. Although such hydrocephalus is in principle preventable, the microbial spectrum that accounts for this disease, and the routes of infection, have never been well characterized.

We have therefore taken a very broad view to this disease, proposing that an unbiased identification of pathogens may be necessary to identify potential causal factors in developing world settings, drawn from the *neonatal septisom*e (*17*) — the assemblage of the pathogens underlying neonatal sepsis in such settings.

Postinfectious hydrocephalus is part of a spectrum of conditions that through activation of the immune system in the brain lead to acquired hydrocephalus. The other major component of post-inflammatory hydrocephalus of infancy is intracerebral hemorrhage of prematurity. Both infection and hemorrhage within the brain lead to hydrocephalus through related inflammatory mechanisms (*18*). PIH is not a disease caused by a single organism, and the use of an unbiased pan-microbial analysis in other regions will likely reveal other organisms as important causes of PIH.One of the problems with utilizing genomic techniques for pathogen detection from nominally sterile body fluids, such as blood and CSF, is that such low-biomass samples are plagued by bacterial DNA contamination from reagents and other sources that can dominate the results (*19*), and substantial effort has addressed both statistical (*20*) and spike-in strategies (*21*) to reduce such effects. By employing case-controls in our study consisting of contemporaneously recruited NPIH patients referred to the same hospital, we were able to rigorously contrast our analysis of infected samples to that of clinically uninfected controls. By replicating our bacterial discovery efforts on differently preserved samples, in independent laboratories with separate regions of 16S, we reproduced convergent results — demonstrating a dominant *Paenibacillus* PIH pathogen in this cohort.

Whether prior analysis of postinfectious hydrocephalus in this region was biased by reagent contamination is not known at this time (*7*), but we implemented several technical strategies to reduce the effect of background contamination (see Supplementary Material). Despite such contamination reduction efforts, the additional use of case-controls was critical to achieve convincing levels of differential abundance significance, along with validation through organism recovery in culture from cases. The organism culture recovery rate was low, potentially due to the use of fresh frozen samples and antibiotic administration prior to patient sampling.

While various *Paenibacillus* spp. have been occasionally isolated from CSF (*22-24*), *P. thiaminolyticus* has not been known to be a virulent pathogen. It was first identified while screening for bacteria in the gut that might contribute to thiamine deficiency in beriberi in Asia (*25*). There is a single case report of indwelling catheter-associated bacteremia in an elderly patient on hemodialysis (*26*). This bacterium produces thiaminase I and II and is adapted to live in low-thiamine environments. Thiamine deficiency is associated with Wernicke’s encephalopathy (*27*), two forms of beriberi (*28*), and polioencephalomalacia-associated brain necrosis in ruminant animals (*29*). In the developing world, thiamine deficiency is common in children (*30*), and is exacerbated by the stress of infection (*31*). Our findings demonstrate that thiamine levels were lower in postinfectious children regardless of infectious etiology, but, consistent with reports in animals (*32*), we did not demonstrate lower thiamine levels specifically due to infection with *P. thiaminolyticus*.

The underpinnings of the organism’s virulence remain uncharacterized at present. We found it difficult to culture this facultatively anaerobic organism from clinical samples. Once established in culture, the organism could be passaged onto aerobic media. We speculate that with a predilection to form calcified loculations and abscesses within the brain, it may be growing anaerobically when sampled, and require initial anaerobic conditions before switching to aerobic metabolism. Alternatively, the lytic properties of the anaerobic broth used successfully in recovery might have released viable organisms from intracellular phagocytosis within white blood cells. In either case, this organism has acquired substantial virulence in comparison with the existing reference strain, as demonstrated by nearly complete lethality of high concentrations of this organism when inoculated into mice in contrast with the reference strain. Supporting this differential virulence are multiple phage insertions into its genome, and other protein coding and copy number variations, that await further characterization.

The expression of differential virulence found in mice was constrained by the rapid lethality. The apparent toxin effect may have precluded replicating the establishment of the brain infections during sepsis which are the hallmark of PIH. Although our findings and animal model do not meet Koch’s postulates for disease causation (*33*), many infectious disease do not meet these criteria (*34*), and in particular PIH can be caused by different organisms. Nevertheless, *P thiaminolyticus* Mbale was the dominant organism present in these PIH cases, correlation with disease severity on brain imaging was substantial, and correlation with central and peripheral WBC counts was strong.

Although this organism appears quite sensitive to common antibiotics, the presence of calcified abscesses will make the penetration of antibiotics to achieve adequate bactericidal concentrations challenging. Ideally, such concentrations are achieved at initial point-of-care treatment for neonatal sepsis prior to infection of the brain. It is possible that within the immune-privileged brain (*35*), inadequate treatment during neonatal sepsis is a substantial factor in the persistent development of brain *P. thiaminolyticus* infection. How co-infection with CMV might affect disease severity in the setting of *P. thiaminolyticus* Mbale infection is unknown at this time.

The typical clinical picture of these PIH cases is that of a ventriculitis without prior meningitis. Whether lumbar puncture in such infants, without meningeal infection, could be diagnostic during their neonatal sepsis evaluation is presently unknown.

Notwithstanding that we find CSF purulence, high levels of matching DNA, and recovered organisms whose DNA matches that in infected cases closely, a full description of causality in this disease may be more complex. Our analysis was limited to detection of active infections. For the regions in which we were working, *in utero* exposure to infections such as malaria is common (*36*). But predisposing *in utero* infections which are no longer active after birth (whether parasitic, viral, or bacterial), would remain undetected in our genomic sampling. Our finding of a substantial viral background infection with CMV cannot distinguish congenital from postnatally acquired viral infection, but the true incidence of *in utero* infections must have been higher than what we could observe through testing several months after birth. In addition, our analysis does not address the heterogeneity in innate immunity (*37*) or nutritional status (*38*) known to be predisposing factors of infection.

The demographics of the *Paenibacillus* infections suggests localization to a circumscribed region in Eastern Uganda. This is a region associated with the north and south banks of Lake Kyoga, and the wetlands along the northern edge of Lake Victoria. It is characterized by large swamps and is a rice-growing region. Whether these infections are influenced by rainfall (*39*), which has been previously observed in PIH without organism identification (*40*), or share similarities with other environmental agents in similar topographies of the developing world (*41*), remains unknown.

Nevertheless, a pan-microbial approach has uncovered the presence of a difficult to grow pathogen not previously known to possess substantial virulence, which appears associated with calcified loculations and brain abscesses in infants, as well as with hydrocephalus following survival from neonatal sepsis. The presence of this organism upon a neurotropic viral background creates a scenario with frequent viral-bacterial coinfections where 6/8 CMV by CSF and 11/27 CMV by blood positive cases were co-infected with *Paenibacillus* spp. Prior studies in Ugandan adults have found that CMV viremia is frequently present in the setting of sepsis and is associated with increased risk of mortality (*42-44*). It has been hypothesized that immune modulation (*45*), rather than direct CMV effects, are responsible for the association of CMV with worse outcomes, especially for tuberculosis and cryptococcal meningitis. It is likely that, in many of these cases, latent CMV reactivates when overwhelming infection by another pathogen alters the immune system’s ability to keep the virus sequestered (*44, 45*). However, due to the ages of our patients, all <90 days of age, a majority of the CMV viremias detected would be expected to be related to primary CMV infection (acquired either congenitally or during early postnatal life) and be more likely to cause, rather than be a cause of, alterations in immune function with related increases in bacterial pathogenesis and increased risk of severe bacterial infection (*46*).

Limitations to this study are important to note. PIH is a syndrome, and although we have identified a novel agent that appears to play a major role in the cases from Eastern Uganda, it remains unclear what other bacterial pathogens may play an important role in other regions of Uganda, or other countries within sub-Saharan Africa and beyond. Whether *Paenibacilus* spp. predisposes to invasion of the nervous system by CMV, or vice versa, remains unknown at present. The non-diagnostic PIH cases are mysteries, in that the ectopic calcifications seen commonly in the *Paenibacillus* positive cases would not have disappeared in the slightly older cohort of non-diagnostic patients – it is possible that they may have harbored different organisms. The prime limitation to our pan-microbial molecular approach seems to have been the age of the patients – only survivors of neonatal sepsis can develop PIH, and our data is consistent with a need to identify the causative organisms as early as possible, ideally during treatment of neonatal sepsis accompanied by serial brain imaging.

Addressing the estimated 160,000 annual cases of PIH generated largely throughout the developing world (*1*), and the larger pool of several million yearly neonatal sepsis cases (*2*), is a critical global public health need. If a pan-microbial approach is required, then the current technology available to achieve this is at present neither readily scalable nor economically sustainable. On the other hand, the expansion of high-technology surgical facilities and pediatric neurosurgical care is also not readily scalable (*47*). Long-term, more sustainable and less expensive technologies to achieve pan-microbial surveillance in such settings, including maternal and environmental sources of infection, will be required to enable more optimal treatment and ultimately prevention of these devastating neonatal infections.

## Methods

### Study design and oversight

The study was conducted at the CURE Children’s Hospital of Uganda (CCHU), a freestanding pediatric neurosurgical hospital in eastern Uganda that serves as a countrywide referral center for patients with hydrocephalus. Infants were eligible for participation in the trial if they were 3 months of age or younger, met criteria for postinfectious or non-postinfectious hydrocephalus (PIH and NPIH respectively), and had a mother who was at least 18 years of age. The study was designed as a waste-fluid study at surgery with verbal consent. Ethics oversight was provided by the CCHU Institutional Review Board, the Mbarara University of Science and Technology Research Ethics Committee, and with oversight of the Ugandan National Council on Science and Technology. The study was approved by the Penn State University Institutional Review Board, and a Materials Transfer Agreement was in place between CCHU and Penn State University. A US Centers for Disease Control permit for the importation of infectious materials covered the transfer of specimens from CCHU to Penn State. An Institutional Biosafety Committee provided oversight of specimen handling at Penn State. A Materials Transfer Agreement between Penn State and Columbia University covered the transport of materials between these research sites.

#### Inclusion criteria for PIH

a) age 3 months or less, b) weight greater than 2.5 Kg, c) no history consistent with hydrocephalus at birth, and either i) a history of febrile illness and/or seizures preceding the onset of clinically apparent hydrocephalus, or ii) alternative findings such as imaging and endoscopic results indicative of prior ventriculitis including septations, loculations, or deposits of debris within the ventricular system (*3*), and d) mothers at least 18 years old to give informed consent.

#### Inclusion criteria for NPIH

a) age 3 months or less, b) weight greater than 2.5 Kg, c) findings of non-infectious origin of hydrocephalus on computed tomography (CT) scan or at endoscopy such as a lesion obstructing the Aqueduct of Sylvius such as tumor or cyst, aneurysm, or cavernous malformation, Dandy-Walker cyst, or other congenital malformation of the nervous system, or d) evidence of hemorrhage as cause of hydrocephalus such as i) bloody CSF and ii) absence of findings consistent with PIH or congenital origin of hydrocephalus, and e) mothers at least 18 years old to give informed consent.

#### Exclusion criteria for hydrocephalus study

a) prior surgery on the nervous system (shunt, third ventriculostomy, or myelomeningocele closure), or b) evidence of communication of nervous system with skin such as meningocele, encephalocele, dermal sinus tract, or fistula.

### Statistical Methods

Continuous demographic variables were evaluated using the non-parametric Wilcoxon rank-sum (two-group comparisons) and Kruskal-Wallis (>2 groups) tests following Shapiro-Wilk’s test for normality, unless otherwise stated. Fisher’s exact test was performed for categorical variables. All tests were two sided unless stated otherwise. Ordinal logistic regression was applied to estimate the proportional odds of CT scan scores between indication, *Paenibacillus* presence or absence, and CMV status. We performed differential abundance analysis of taxa abundances between PIH and NPIH groups and accounted for multiple testing leveraging false discovery rate analysis (*48*).

### Brain image evaluation

Preoperative CT scans were independently scored, blindly with respect to diagnosis, by two board certified neurosurgeons who have considerable experience with infant hydrocephalus (BCW, SJS). One point was assigned for each of four possible findings: fluid loculations, debris within fluid spaces, ectopic calcification within the brain parenchyma, and abscess formation. Discrepancies between scoring were then resolved to a consensus agreement.

### Sample collection and storage

Blood was sampled with aseptic technique at the time of surgery, either at the time of catheter placement for an intravenous line or during venipuncture for routine laboratory testing. CSF was obtained at the time of initial surgery. Many of the PIH cases were treated for abscess formation once or twice, prior to a definitive surgical procedure to address the hydrocephalus (endoscopic third ventriculostomy with choroid plexus cauterization, or ventriculoperitoneal shunt insertion). Initial surgery might not employ an endoscope if the primary goal was aspiration and irrigation of purulent material from loculated fluid collections or frank abscesses. These initial procedures might also include endoscopic lateral terminalis and septum pellucidum fenestrations. The endoscope was nearly universally used in all other cases, such as NPIH, as is our standard practice at this surgical site. Samples of blood and spinal fluid were divided into aliquots for fresh freezing or placement into DNA/RNA preservative (DNA/RNA Shield, Zymo Corporation), and specimens were frozen either at -80°C or placed in a liquid nitrogen Dewar or dry shipper. They were kept frozen through transport to the US for further analysis.

### Ribosomal RNA (rRNA) 16S gene sequencing

For characterization of bacterial species we performed 16S rDNA amplicon sequencing. Separate CSF samples were sequenced at two different laboratories. Independent approaches were applied to limit background amplification of contaminants and decontaminate reagents (see Supplementary Materials). At one laboratory, 16S amplicon sequencing of the V1-V4 region was performed on fresh frozen samples using Sanger sequencing. Further next generation amplicon metabarcode sequencing was performed on V4 for microbial background characterization, and *Paenibacillus* genus-specific qPCR for quantification was performed. At the other laboratory, a primer extension technique for 16S amplicon next generation sequencing of the V1-V2 region on DNA/RNA preserved samples was performed. Utilizing results from these two laboratories, 16S amplicon (regions V1-V2 and V4) reads sequenced from fresh frozen CSF and preservative samples were clustered at 97% similarity (*49*). For downstream analyses we accounted for sequencing variability using cumulative sum scaling normalized taxa abundances (*50*). A primer table is given in Table S1.

### Targeted pathogen gene testing

Targeted polymerase chain reaction (PCR) was performed in an attempt to detect the presence of Zika virus, chikungunya virus, human papilloma virus, parvovirus B19, toxoplasmosis, trypanosomiasis, malaria, and fungi (Table S1).

### Virus detection

A broad screen for viral presence was performed in two different ways: VirCapSeq oligomer concentration (*10*) and total RNA sequencing analysis. For the viruses that appeared abundant in either PIH or NPIH, PCR confirmation was performed.

### Metabolites

Thiamine diphosphate was quantified in fresh frozen blood using high-performance liquid chromatography with tandem mass spectrometry (LC/MS/MS) at Mayo Clinic Laboratories.

### Microbiology

In an attempt to culture the putative pathogen, 100 fresh frozen CSF samples were subjected to 6 different media outlined in Table S8. If colonies grew on solid media, gram stain and matrix-assisted laser desorption/ionization time-of-flight (MALDI-TOF) were performed to characterize the organism. For isolates identified as *Paenibacillus*, antibiotic sensitivity, biochemical testing, and whole genome sequencing were performed.

### Genome assembly

A hybrid method was utilized to reconstruct the genome of a *P. thiaminolyticus* isolate, combining short read sequencing, optical mapping (Bionano Genomics), and nanopore long contiguous sequencing (MinION, Oxford Nanopore Technologies). From the resulting whole genome sequences and optical mapping assembly a hybrid scaffold was generated (Bionano Hybrid Scaffold v1025201). For *P. amylolyticus* and *Paenibacillus* spp. isolates, and reference type strain *P. thiaminolyticus* NRRL B-4156 (Agricultural Research Service Culture Collection https://nrrl.ncaur.usda.gov/cgi-bin/usda/prokaryote/report.html?nrrlcodes=B-4156), only short read and nanopore sequencing were utilized for assembly.

### Animal model virulence testing

All animal experiments were performed with oversight by the Penn State Institutional Animal Care and Use Committee, and with Institutional Biosafety Committee approval at biosafety level 2 (BSL2). Virulence testing was performed on weanling P21-P28 C57BL/6J mice using up to 10^9^ colony forming units suspended in 100 μL saline, or saline only, injected into the peritoneum. Bacteria for injection were thawed and subcultured prior to each inoculation and quantified using standard colony forming unit methods (Supplementary Material). Animals were humanely euthanized with CO_2_ if they developed altered or depressed mentation, or lost more than 20% of their body weight. A full complement of tissues were collected from each mouse following the guidelines set forth by international veterinary toxicology interest groups (*51-53*). Tissues were preserved in 10% neutral buffered formalin, embedded in paraffin blocks, cut into 3 μm sections, and stained with hematoxylin and eosin for analysis. All organs were evaluated by a veterinary pathologist (HA).

## Data Availability

The assembled genome of Paenibacillus thiaminolyticus Mbale was deposited in GenBank with Accession CP041404. Sequencing data for bacterial 16S DNA, in silico host-depleted mRNA, and VirCapSeq data, along with sample metadata are available at the NCBI archive under project ID PRJNA605220. All custom code utilized in this study will be made available to investigators upon request. All data associated with this study are in the paper or supplementary materials.

## Acknowledgments

We are grateful for the discussions and support of N. D. Olson, A. Patterson, Q. Liu, M. Mwebingwa, M. Poss, V. Kapur, D. Craft, and the technical assistance of K. Moran.

## Author contributions

The study was designed by J.K., and S.J.S. The manuscript was written by J.N.P, B.L.W., C.H., and S.J.S. The data analysis was performed by J.N.P., B.L.W., C.H., N.M., L.Z., S.M.K., M.P. Bacteriology was carried out by D.S.S.W., B.V.B., and J.B., and advised by M.Y.G. and M.A. Virology was performed by N.M. Wet lab technical analysis was performed by C.H., M.C-R., C.G., and J.N. Animal experiments were carried out by P.S., and histology performed by H.A. Independent laboratories were supervised by W.I.L. and J.B. Clinical work was performed by J.M., F.B., E.M-K., R.M., E.K., J.M., P.O-O., J.O., K.B., P.S. Project management was performed by S.S., E.M-K., J.K., E.K., K.S., A.D.W., M.G., T.W., W.I.L, J.B., and S.J.S. Bioinformatics was performed by J.N.P., L.Z., F.R., and J.Q. Neurosurgical consultation was performed by A.V.K., D.D.L., B.C.W., and S.J.S. Infectious disease consultation was provided by L.B. and J.E.E. Computer support by B.N.K. Immunology consultation was performed by S.U.M. and M.H. CSF proteomics was performed by A.M.I., R.T., and D.D.L. Statistical analysis was provided by J.N.P., M.H., and X.L., and geographical mapping performed by A.J.W. and P.S. All authors contributed to editing the manuscript.

## Data and materials availability

The assembled genome of Paenibacillus thiaminolyticus Mbale was deposited in GenBank with Accession CP041404. Sequencing data for bacterial 16S DNA, in silico host-depleted mRNA, and VirCapSeq data, along with sample metadata are available at the NCBI archive under project ID PRJNA605220. There was a Materials Transfer Agreement between the provider CURE Children’s Hospital of Uganda and recipient Penn State University, where Penn State University retains rights to the derivatives, *Paenibacillus thiaminolyticus* Mbale and for the Mbarara University of Science and Technology, CURE Children’s Hospital of Uganda, and Penn State University to own any new products discovered through the use of the materials, *Paenibacillus thiaminolyticus* Mbale. There was a Materials Transfer Agreement between the provider Penn State University and recipient Columbia University, for Penn State University to retain ownership of the materials, *Paenibacillus thiaminolyticus* Mbale, and joint ownership of modifications. All custom code utilized in this study will be made available to investigators upon request. All data associated with this study are in the paper or supplementary materials.

## List of the Supplementary Materials

Materials and Methods

Figure S1: CT Scans.

Figure S2: Spatial Statistics.

Figure S3: Phylogenetic tree for 16S V1-V4 sequences.

Figure S4: Further analysis of the microbial 16S community.

Figure S5: OTU Heatmaps.

Figure S6: qPCR vs Age, CT Score, and Cell Counts.

Figure S7: CT Score Frequencies.

Table S1: Primer Table.

Table S2: Fresh Frozen PCR and Sequencing.

Table S3: Differential Abundance.

Table S4: Viral Results.

Table S5: Demographics with and without Paenibacillus.

Table S6: Paenibacillus vs CMV.

Table S7: CT Score vs CMV.

Table S8: Culture Media.

Table S9: Culture Results.

Table S10: Biochemical testing of P. thiaminolyticus.

Table S11: Antibiotic Resistance.

Table S12: Virulence Testing.

References (*56-105*)

## Supplementary Material

## Microbial Characterization

We describe in detail the wet lab and computational methods for microbial characterization as well as *P. thiaminolyticus* sequencing and assembly. 16S rRNA gene amplicon sequencing was employed to initially characterize the bacterial flora present in patients’ CSF for the characterization of dominant organisms and potential pathogen detection. 16S rRNA amplicon sequencing is commonly used for microbial community characterization, including differential abundance analysis. However, a limitation to 16S rRNA amplicon sequencing is a lack of taxonomic resolution, where organisms are often only identifiable to the genus or family level.

Through targeted amplicon sequencing we were able to characterize the flora and evaluate abundant and variable taxa within postinfectious hydrocephalus (PIH) cases and those differentially abundant between PIH cases and non-postinfectious hydrocephalus (NPIH) controls. Noticeably, we identified a *Paenibacillus* taxa both dominant and differentially abundant in PIH cases. Following detection, we isolated the organism, performed quantification using qPCR, biochemical testing, matrix-assisted laser desorption/ionization time-of-flight (MALDI-TOF) mass spectrometry, and marker gene taxonomy placement following genome assembly. All primers for these methods and their appropriate references are listed in Table S1.

## Extraction, Amplification, and Sequencing

Below we present the extraction, amplification, and sequencing details for the fresh frozen and preserved CSF samples.

### Fresh Frozen CSF Samples

#### Nucleic Acid Extraction of Fresh Frozen CSF Samples

Prior to extraction, all tubes, columns, 0.1 mm and 0.5 mm glass beads (MoBio, CA, USA) and 1 mL extraction reagent aliquots were UV-irradiated twice at a distance of one inch from UV bulbs and at a setting of 3000 × 100 µJ/cm^2^ in a spectroLinker XL-1500 UV crosslinker (Spectronics Corporation, NY, USA). All of the kit extraction reagents (liquid) were aliquoted into 2 mL tubes in a UV hood at a volume not exceeding 1 mL and were UV-irradiated as above. To control for any remaining contamination, ten extraction reagent controls (blank extractions, negative extraction controls) were also included. Nucleic acid from fresh frozen CSF was extracted using the QIAgen AllPrep DNA/RNA kit (QIAgen, MD, USA), with some modifications. Before extraction, 1.5 mL of CSF was first centrifuged at 5600 rpm for 10 minutes at 4°C to pellet all cells and 500 μL of supernatant was removed for viral analyses. To the remaining 1.0 mL of CSF used for bacterial and fungal analysis, Buffer RLT with 0.01% ß-mercaptoethanol (600 mL and 700 ml, respectively) and 3 μL of DX reagents (QIAgen, MD, USA) were added along with 0.1 mm and 0.5 mm glass beads. Samples were disrupted in a Tissue Lyser (QIAgen, USA) for five minutes at 30 Hz followed by proteinase K digestion (6 mAU, Novagen, USA) done at 55°C for 30 minutes then all remaining steps were performed according to the manufacturer’s protocol. RNA and DNA concentration and purity were measured with a NanoDrop ND-100 spectrophotometer (NanoDrop Technologies, DE, USA) and stored at -80°C.

#### Bacterial V1-V4 16s rRNA gene and Fungal ITS2 amplification

For PCR of both bacterial 16S rRNA and fungal ITS from fresh frozen CSF, consumables and prepared master mix (without primers) were UV-irradiated in a SpectroLinker XL-1500 UV crosslinker (3000 × 100 μj/cm^2^) prior to PCR setup. Once the prepared master mix was UV-treated, primers were added and a restriction digest was performed using Sau3AI at 37°C for 30 minutes then chilled on ice for 5 minutes prior to template DNA being added. The following controls were used for all reactions: ten negative extraction control, three negative PCR controls (no template), and three positive PCR controls (DNA from human stool for bacteria or soil for fungi) included. Primers for 16S V1-V4 amplification were 27F.1 and 805R and primers for ITS2 amplification were ITS3F and ITS4R. Each 20 μL PCR reaction consisted of 1X Accuprime Buffer II, 0.75 units of Accuprime Taq DNA Polymerase High Fidelity (Life Technologies, Thermo Fisher, USA), 2.5 units of Sau3AI restriction enzyme, 400 nM of each primer and 5 μL of the extracted DNA. For 16S rRNA amplification, cycling conditions were as follows: 95°C for 5 min, followed by 40 cycles of 95°C for 30 s, 52°C for 1 min, 72°C for 1.5 min, followed by a final elongation step at 72°C for 7 min. For ITS2, cycling conditions were the same except the extension time at 72°C was 1 minute during cycling and the final extension at 72°C was 5 minutes.

PCR products were run on a 1% agarose gel stained with GelGreen (Biotium, CA, USA). None of the extraction reagent controls or PCR reagent controls produced any signal on agarose gels. Visible bands were excised from the gel and purified using the QIAgen Gel Extraction kit according to manufacturer’s guidelines. Purified amplification products were ligated into the pGEM-T Easy vector (Promega Corporation, WI, USA) and inserts (at least 5 clones per sample) were sequenced by Sanger sequencing (Genewiz, NJ, USA).

#### V4 Illumina Sequencing of bacterial 16s rDNA

DNA from fresh frozen CSF samples were amplified with composite barcoded primers targeting the V4 region of the bacterial 16S rRNA gene to generate libraries for Illumina MiSeq sequencing according to the Earth Microbiome Project standard protocol (http://www.earthmicrobiome.org/emp-standard-protocols/16s/) with minor modifications (*56*). The same decontamination techniques were employed as described above, except that two restriction enzymes were used to decontaminate PCR reagents in each reaction, 2 units each of Sau3AI and AciI. The same decontamination techniques were employed as described in full length 16S and ITS2 PCR amplifications techniques described above. The PCR cycling conditions were as follows: 94° C for 5 min, 40 cycles of 94°C for 20 s, 53°C for 25 s, 68°C for 45 s and a final extension at 68°C for 10 min. Amplification products (2 μl) were run on a 1% agarose gel stained with GelGreen (Biotium, CA, USA) to verify amplification and negative reactions were verified on a BioAnalyzer. None of the extraction reagent controls or PCR reagent controls produced any signal on agarose gels or BioAnalyzer. PCR products were further purified using Ampure magnetic purification beads (Beckman Coulter Life Sciences). Ampure purified products were quantified with the Quanti-iT PicoGreen dsDNA Assay Kit (Invitrogen). Equimolar ratios of each sample were combined to create DNA pools of barcoded libraries for sequencing on an Illumina MiSeq.

#### Real-time Quantitative PCR (qPCR) of *Paenibacillus* spp. from CSF

*Paenibacillus* species specific qPCR was performed targeting 16S rDNA as previously described with modifications (https://www.sciencedirect.com/science/article/pii/S0022201111001108?via%3Dihub). For absolute quantitation, DNA standards for *Paenibacillus* were constructed by PCR amplifying DNA obtained directly from *Paenibacillus*-positive CSF samples from this study. Purified amplification products were ligated into the pGEM-T Easy vector (Promega Corporation, WI, USA) and confirmed by Sanger sequencing. After verifying the plasmid insert sequence, 10-fold serial dilutions of plasmids were generated, ranging from 5 × 10^6^ to 5 copies and were spiked with salmon sperm DNA (2.5 ng/μL). PCR efficiency for plasmid standards was above 90% with correlation coefficients ranging from 0.996 to 1, and sensitivity was 5 copies. Each 25 μL real-time PCR reaction consisted of 1x TaqMan Universal PCR Master Mix (Applied Biosystems, Thermo Fisher Scientific, USA), 5 μL of fresh frozen CSF DNA and primers (PaeniF and PaeniR,250 nM) and a probe (PaeniProbe, 300 nM). An ABI Step-One Plus Real-time PCR system (Applied Biosystems) was used to perform all real-time PCRs using the following cycling conditions: initial denaturation at 95°C for 15 minutes followed by 45 cycles at 95°C for 15 sec and 60°C for 1.5 min. All samples were run in duplicate for each assay, and average copy number was calculated from duplicate reactions. Final results are presented as total *Paenibacillus* spp. 16S rDNA copies per mL of CSF.

### Preserved CSF Samples

#### Nucleic Acid Extraction of Preserved CSF Samples

Nucleic acid extractions on DNA/RNA Shield (Zymo, CA, USA) preserved samples were performed with reagents and consumables UV-treated in a PCR Workstation for 15 minutes. Each batch of extraction included a water control in which 500 μL of elution buffer was put through all extraction steps. DNA was extracted from shield samples using 500 μL of mixed sample using ZymoBIOMICS DNA Miniprep Kit (Zymo, CA, USA) with bead lysis using 0.15 mm and 0.5 mm ZrOBO beads in Bullet Blender (Next Advance, NY, USA) at high speed for 5 minutes. After lysis, the manufacturer’s protocol was followed with proteinase K digestion and two elutions of 50 μL with elution buffer heated to 65°C. RNA was extracted from shield samples using TRIzol™ LS Reagent and Direct-zol™ RNA MiniPrep Plus kit. Samples were processed with two aliquots of 250 μL mixed with 750 μL of TRIzol™ LS Reagent. After bead lysis, 200 μL of chloroform was added, incubated at room temperature for 15 minutes then centrifuged at 12,000g for 15 minutes at 4°C. Once separated the aqueous layers from the two aliquots were put into the same tube. Then each organic layer was re-suspended in 400 μL of water and centrifuged again for 5 minutes. The 1.6 mL of aqueous phases were combined with 1.6 mL of 100% ethanol and put through the Zymo-Spin™ IC Column twice and proceeded per manufacturer’s protocol.

#### Bacterial 16S rRNA Library Prep and Sequencing with Primer-extension PCR

Primer-extension PCR (PE-PCR) of the 16S rRNA V1-V2 region using primer 336R with M13 and 27F with previously described techniques by (*57*) with a few modifications. To control temperature appropriately the initial annealing step was done on the the Hybex microsample incubator (Scigene, CA, USA). To avoid any additional contamination, MolTaq 16S Mastermix (Molzym GmbH & Co Kg, Germany) was used for amplification following manufacturer’s protocol with the following optimized PCR cycling conditions: 35 cycles of 15 seconds at 95°C and 2 minutes at 60°C. All of the PE-PCR product was put into a 1x AMpure XP (Beckman Coulter) cleanup and eluted in 50 μL of 10 mM Tris pH 8. Library preparation was performed using the Hyper Prep Kit (KAPA Biosystems, USA) according to manufacturer’s protocol, adjusting adapter concentration based on total DNA input. Libraries were subsequently enriched with 7 cycles of PCR using the Library Amplification Module (KAPA Biosystems, USA). Libraries were quantified using Agilent Bioanalyzer DNA 1000 chip, pooled to an appropriate concentration and another 1x clean-up was performed. Sequencing was performed per the manufacturer’s protocol on Illumina’s Miseq using the 600 cycle v3 kit at 9 pM and 7% phiX with an aimed depth at 500k reads/sample.

## 16S Analysis

Below we present the analytical overview of the 16S analyses on our fresh frozen and preserved CSF samples. We first highlight the *in silico* comparison of V1-V2 and V4 amplicon regions for potential characterization based on Greengenes databases. Afterwards, we present our process for generating Operational Taxonomic Unit (OTU) count tables, annotation, differential abundance analysis, and comparison of diversity by sample type.

### *In silico* V1-V2 and V4 amplicon region characterization

We define taxonomic resolution as the ability to differentiate between groups within a taxonomic level, for example differentiating between species within a genus. Taxonomic resolution can vary by clade and amplicon regions, though the extent to which it varies is not well characterized. Below in Section Comparison of V1-V2 and V4 we highlight the differential potential to speciate *Paenibacillus* taxa followed by the comparison of diversity levels and relationship to CSF cell count.

We utilized metagenomeFeatures (*58*) and the MgDb annotation packages (*59*) to characterize taxonomic resolution for a specific clade and amplicon region, specifically for the *Paenibacillus* genus and V1-V2 and V4 regions using the Greengenes 13.5, greengenes13.5MgDb.us (http://bioconductor.org/packages/release/data/annotation/html/greengenes13.5MgDb.html). The number of sequences assigned to specific *Paenibacillus* species, range from 199 for *Paenibacillus amylolyticus* to 2 for *Paenibacillus illinoisensis*. Sequences only classified to the genus level, “Unassigned”, is the most abundant group, 2308.

16S rRNA amplicon sequencing taxonomic resolution for *Paenibacillus* species were compared by calculating within and between species pairwise distances. In order to differentiate between species, the pairwise distances within species amplicon sequences must be less than the between species distances. Additionally, the difference in amplicon sequence pairwise distances between and within species must be greater than the sequencing error rate to detect the difference. For our taxonomic resolution analysis we use *in silico* PCR to extract the V1-V2 and V4 regions of the 16S rRNA sequences. We generated a pairwise distance matrix for the two regions and compared the within and between species pairwise distances using DECPIHER (https://decipher.sanger.ac.uk/). For our *in silico* PCR we used the bacterial portion of the PCR primers from Table S1 as: For the V1-V2 and V4 analyses, only sequences with the appropriate expected amplicon lengths are extracted (318 bp and 252 bp for the V1-V2 and V4 regions respectively). The overall pairwise distance is greater between species than within species for the *Paenibacillus* genus.

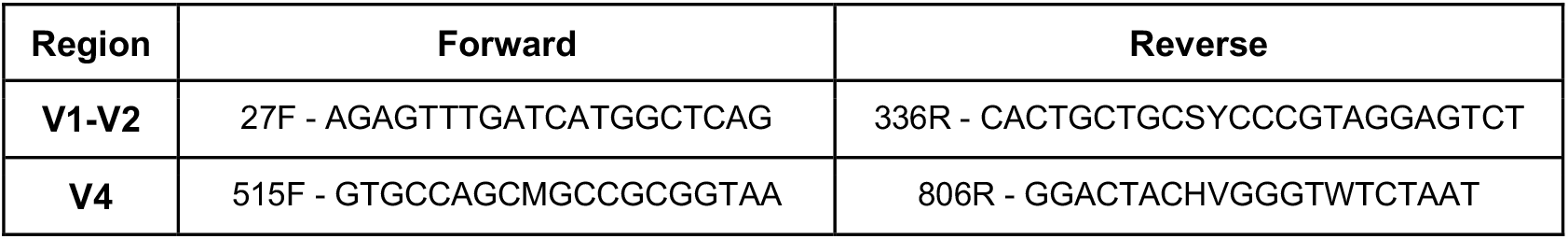

### OTU Matrix Generation

An open-reference OTU matrix was generated using Qiime version 1.9.1 (*60*) for all downstream analyses. In the process, reads were clustered using a 97% similarity cutoff with uclust version 1.2.22 and aligned using PyNAST v1.2.2 against a modified core Greengenes database vgg_13_8 (*61*) that included 16S references for three strains successfully isolated from clinical samples (see below). *De novo* OTUs were generated for unaligned reads. OTU centers were filtered due to chimeric detection using ChimeraSlayer (via microbiomeutil_r20110519) (*62*). OTU centers were assigned a taxonomic annotation using RDP Classifier v2.0.2 (*63*). Further taxonomic assignment for *Paenibacillus* of unannotated OTUs consisted of annotating centers against the NR database (updated with NCBI on Oct. 3rd, 2018) with BLAST.

#### 16S OTU Processing from Fresh Frozen CSF Samples

Two thousand one hundred and five (2,105) OTUs were generated from the process described above. Of these, 658 passed chimera checking, were detected in more than five samples or represented at least 20 sequences in a single sample, and were included in further analysis. The number of OTUs per sample ranged from 51 to 296, with a median of 125 and an average of 137. The mean OTU size was 2328, ranging from 5 (by definition) to 736,735 (with median OTU size = 58 sequences). Among these, approximately 56% of OTU centers were annotated to the genus level using the Greengenes database, while others were annotated at family or higher levels.

#### 16S OTU Processing from Preserved CSF Samples

Five thousand eighty-eight (5,088) OTUs were generated from targeted 16S rRNA amplicon sequencing. Of these, 1,767 passed chimera checking, were detected in more than five samples or represented at least 20 sequences in a single sample, and were included in further analysis. The number of OTUs per sample ranged from 22 to 429, with a median of 140 and an average of 156. The mean OTU size was 3,331, ranging from 5 (by definition) to 2,071,935 (with median OTU size = 126 sequences). Among these, approximately 66% of OTU centers were annotated to the genus level using the Greengenes database, while others were annotated at family or higher levels.

### Characterization of samples and clinical association

#### Microbial Overview

In processing samples, we prepared bar plots of the most abundant organisms present. Caution in interpretation of the background microbial diversity and community is necessary as CSF samples are low-biomass samples known to yield PCR and other sequencing artifacts (*64*). That being said, a number of patient samples yielded many reads from single taxa not known to associate with sequencing artifacts. Fig. 2C and 2D and Tables S3 and S4 highlight the most abundant organisms from both preservative CSF (V1-V2 amplicon sequencing) and fresh frozen (V4 amplicon sequencing) samples, respectively.

#### Similarity and diversity analysis

The background microbial community and associations with clinical samples for the fresh frozen and preserved samples were compared. In particular, we compared *Paenibacillus* abundances for matching pairs of samples calculated by aggregating annotated OTU normalized counts and log-transforming, highlighting the similarity in abundance for various OTUs. We observed similar clustering of patients by *Paenibacillus* spp. V1-V2 and V4 OTUs (Fig. 2E, Fig. S5).

Overall, diversity was lower in V4 samples potentially due to both the shorter V4 region and a different strategy of reagent contamination reduction, resulting in fewer OTUs. Diversity was inversely correlated with CSF cell count with the predominance of a single dominant organism in those samples as would be expected (Fig. S4B). The detection of *Paenibacillus* spp. presence in PIH samples was observed across multiple batches; in particular, sequencing and extraction batching was performed with a mix of PIH and NPIH samples to control for any artifacts.

#### Differential abundance analysis

Differential abundance and presence/absence analysis was performed to compare PIH and NPIH taxa abundance and presence. For differential abundance analysis normalized counts were aggregated to the genera level and a log-normal regression model was applied using limma (https://bioconductor.org/packages/release/bioc/html/limma.html) with abundance as the outcome variable and indication status as the independent variable. Counts were normalized using cumulative sum scaling and aggregated to the genera clade (*65*). For differential presence/absence testing we applied Fisher’s exact test on the presence/absence of each genus as defined by the presence of a positive count, also implemented within MetagenomeSeq (https://bioconductor.org/packages/release/bioc/html/metagenomeSeq.html). To account for multiple testing Benjamini and Hochberg’s false discovery rate was applied (*66*).

## Bacterial Isolate Whole Genome Sequencing and Assembly

### DNA Sequencing

From cultured bacteria, high molecular weight DNA was prepared for optical mapping and both short and long read whole genome sequencing per the Prep Cell Culture DNA Isolation Protocol including the necessary Lysozyme digestion for bacterial cells (Bionano Genomics, CA, USA). PCR-free short read libraries were generated from 1 ug of E220 Focused Ultrasonicator (Covaris, USA) fragmented DNA, average insert size 400 bp with Hyper Prep Kit (KAPA Biosystems, USA). Final library pools were diluted to 10 pM, combined with 15% phiX, and sequenced to 10 million reads each with a MiSeq v3 600 cycle reagents. Long read sequencing library prep was performed using Hyper Prep kit (Kapa Biosystems, USA) and 1D Sequencing kit (Oxford Nanopore Technologies, UK) following the manufacturer’s protocol. Sequencing was done using the MinION with the SQK-LSK108 SpotON flow cell following manufacturers protocol.

### Bacterial Genome Assembly

Reads were aligned to hg38 using Bowtie 2 v2.3.4.3 to remove any spurious human reads. SPAdes v1.3.11 (*67*) was used for de novo scaffold generation using Illumina short read sequencing. To correct reads with scaffolds generated from Illumina short reads Pilon was employed (*68*). Basecalling and reads from MinIon were preprocessed using Albacore (Oxford Nanopore Technologies, https://nanoporetech.com/) Canu was used to assemble the MinIon long-reads (*69*). To generate the hybrid scaffold from optical mapping and corrected MinIon sequencing scaffolds the following Bionano Software was used: Bionano Access v1.3.0, Bionano Tools v1.3.8041.8044 and Hybrid Scaffold v10252018.

## *Paenibacillus thiaminolyticus* Identification and Characterization

Below we present the various methodologies applied for characterization. We initially characterize the V1-V4 16S gene Sanger sequencing of identified *Paenibacillus* cases from fresh frozen CSF and generate a tree for relative relationship. Then we grew several isolates in culture, performed whole genome sequencing described earlier, and performed taxonomic placement using genomic marker genes.

### Phylogenetic Analyses of V1-V4 from Paenibacillus

*Paenibacillus* spp. V1-V4 16S rDNA sequences from fresh frozen CSF were phylogenetically compared to each other. Analyses were conducted using MEGA7 (*70*). Sequence alignments were based on representative 16S rDNA sequences obtained from PCR amplification using 27F.1 and 805R primers and subcloning (as described above) from CSF of individual PIH patients in this study. Primer sequences were trimmed from the sequences. 16S rDNA sequences from *Paenibacillus* isolates and related species in Genbank were trimmed to the length of the sequences obtained from the CSF of PIH patients.

The evolutionary history was inferred using the Neighbor-Joining method (*71*). The optimal tree with the sum of branch length = 1.20929307 is shown in Fig. S3. The percentage of replicate trees in which the associated taxa clustered together in the bootstrap test (500 replicates) are shown next to the branches (*72*). The tree is drawn to scale, with branch lengths in the same units as those of the evolutionary distances used to infer the phylogenetic tree. The evolutionary distances were computed using the Kimura 2-parameter method (*73*), and are in the units of the number of base substitutions per site. The analysis involved 39 nucleotide sequences. All ambiguous positions were removed for each sequence pair. There were a total of 750 positions in the final dataset.

### Characterization for Organisms Recovered through Culture

All 100 CSF samples were blindly cultured using six different media with varying inoculum and conditions listed in Table S8. If colonies grew on solid media, Gram stain and MALDI-TOF were performed to characterize the organism. For MALDI-TOF genus and species identification cutoff values were 1.7 and 2.0 respectively. If the BACTEC™ FX instrument flagged positive liquid culture bottles, a Gram stain and sub-cultures were performed as follows: 1) Positive PEDS bottles were subcultured onto CHOC and incubated at 37°C, 5% CO2 for 5 days 2) Positive ANA bottles were subcultured onto CHOC, anaerobic blood agar plate (ABAP) and incubated at 37°C, for 5 days in 5% CO2 (CHOC) or anaerobically (ABAP, Anoxomat System). For negative BACTEC™ liquid culture bottles gram stain was performed and blind cultures were done as follows: 1) For negative PEDSs bottles, subculture was done on CHOC/BAP and incubated at 37°C, 5% CO2 for 5 days 2) For negative ANA bottles, subculture was done on CHOC, anaerobic blood agar plate (ABAP) and incubated at 37°C, for 5 days in 5% CO2 (CHOC) or anaerobically (ABAP, Anoxoimat Sysytem). Biochemical testing was performed using API 50 CH strip following manufacturers protocol. MALDI-TOF was used for initial screening of clinical isolates, but whole genome sequencing was used for definitive classification of all *Paenibacillus* isolates.

Six CSF samples were positive for bacterial growth during initial incubation (within 14 days of incubation), and the results summarized in Table S9. In addition, there were 4 positive blind cultures. Two of the blind ANA cultures were positive for *Propionibacterium acnes*, 2018 and 2081, with MALDI-TOF scores of 2.28 and 2.26 respectively. A third ANA blind culture was positive and identified as *Paenibacillus* species with a MALDI-TOF score of 1.83. One of the blind PEDS cultures was positive and identified as *Bacillus infantis* with MALDI-TOF of 2.25. The API 50 CH system with API CHB/E media (BioMerieux) was used for biochemical characterization of the organisms. After a 48h incubation, both 2033 and NRRL B-4156 were identified as *P. thiaminolyticus* at a 99.9% confidence level (Table S10).

### Taxonomic Placement using Genomic Marker Genes

*Paenibacillus* genomes, n=438, were downloaded from the Genbank ftp website (using the file ftp://ftp.ncbi.nlm.nih.gov/genomes/genbank/assembly_summary_genbank.txt, November 2018). The 16S rRNA sequences of the 438 genomes were compared to the SILVA database (v132) (*74*) to filter the non-*Paenibacillus* genomes, by removing genomes with 16S rRNA identity to *Paenibacillus* lower than 97% on at least 300 nucleotide alignment length using BLASTN (*75*).

Of the 438 genomes, 361 genomes were retained, due to fragmented 16S rRNA or not matching *Paenibacillus* taxa, and added with the pool of the newly sequenced clinical strains 2033, 2006, 2009 and the genome *Saccharibacillus sacchari DSM 19268* used to root the tree. For comparison, proteins in each genome were predicted using Prodigal (*76*), and 40 conserved phylogenetic marker proteins were extracted for each genome using the standalone software fetchMG v1.0 (*77*) and aligned using MUSCLE v3.8.425 (*78*). Duplicated markers in the genome were not included to reduce possible contaminant bias, and all genomes with fewer than 30 markers detected were not included in the tree. The aligned markers were then concatenated to produce a single chimeric amino acid sequence.

A phylogenetic tree was generated (Fig. 5C) using these chimeric concatenated sequences using FastTree v2.1.10 (*79*) using the options *-gamma -pseudo -spr 4 -mlacc 3 - slownni* to account for partial markers and increase exhaustiveness. The Newick FastTree file was then visualized using iTOL (*80*).

The *Paenibacillus* isolation origins were manually explored using the NCBI and PATRIC website (*81*) and classified into 8 categories (animal, environment, human, milk, water, soil, plant, other) and considering rhizosphere isolation as a plant category. The isolation origin was then stored and structured into an iTOL TREE_COLORS file and display in the iTOL tree.

## Bacterial Antibiotic Sensitivity Testing

A bacterial inoculum was prepared by emulsifying several well-isolated colonies from an overnight bacterial growth on agar plate into 3 ml of sterile water to achieve the turbidity of 0.5 McFarland standard. *S. aureus* ATCC 29213 strain was used as quality control. A sterile swab soaked with inoculum suspension was used to inoculate a Mueller Hinton Agar with Blood plate (15×150mm). Plates were allowed to dry for 5 min to absorb excess moisture. Etest strips with antibiotics were placed on the plate within an equidistant pattern using a template. Plates were incubated at 37°C, 5% CO2 for 20 hours and mean inhibitory concentration (MIC) read and interpreted according to Clinical & Laboratory Standards Institute guidelines. The antibiotic resistance results are summarized in Table S11.

## Animal Model Virulence Testing

For preparations of the bacterial culture for injection, bacteria grew for 18-24h on Trypticase Soy Agar (TSA II) with 5% sheep blood (BD) (SBA) plates, and were then suspended, washed once, and resuspended in sterile saline. Then, 10-fold serial dilutions were prepared in sterile saline, and 100 μL were plated on SBA. After 18-24h, growth was assessed and CFU/ml in the original inoculum were estimated from the colony counts on the first plate on which 10-100 colonies grew.

## Viral and Parasite Characterization

We describe in detail the wet lab and computational methods for viral characterization.

### Parasite and viral PCR

Detection of Zika virus (ZIKV), dengue virus (DENV), chikungunya virus (CHIKV), West Nile virus (WNV) and human RNAse P gene was done using CII-ArboViroPlex rRT-PCR assay (*82*).

For cDNA synthesis of Trypanosoma brucei, Trypanosoma cruzi, Parvovirus, Enterovirus, Plasmodium, and Salmonella typhi, Superscript III (Invitrogen cat# 18080044) was used following the manufacturer’s protocol. The cDNA was used with the same assays done with DNA listed below.

For testing of Plasmodium, Salmonella Typhi, Toxoplasmosis, Trypanosoma brucei, Trypanosoma cruzi, Parvovirus, and Enterovirus conventional PCR with DNA input was used using the primers and references in Table S1. Amplitaq Gold master mix (Life Technologies, cat #4398881) was used per the manufacturer’s protocol with 25 µl volume with 1 µl of primers (15 µM) and 1 µl of cDNA or DNA. For all cycling conditions, they were either unchanged from previous work referenced (Table S1) or they had the following modifications:

1. Parvovirus at 95°C for 15 minutes, followed by 40 cycles of 95°C for 30 seconds, 56°C for 30 seconds, 72°C for 30 seconds followed by 72°C for 5 minutes
2. Enterovirus at 95°C for 15 minutes, followed by 15 cycles at 95°C for 30 seconds, 65°C for 30 seconds (minus 1 degree per cycle), and 72°C for 30 seconds, followed by 30 cycles of 95°C for 30 seconds, 50°C for 30 seconds, 72°C for 30 seconds followed by 72°C for 5 minutes.

In addition to conventional PCR, a TaqMan qPCR assay was used to detect Salmonella typhi and Plasmodium independently. Taqman 2x Universal PCR MasterMix (Thermofisher Cat# 4304437) was used in 25 µl reactions with 5 µl of cDNA and 0.5 µl of 15 µM primers and probe. Thermal cycling and detection was done on the CFX96 Real-Time PCR Detection System (Bio-Rad). Following cycling conditions were used for amplification: 95°C for 2 min, followed by 45 cycles of qPCR at 95°C for 15 s and 60°C for 1 min. Fluorescence signal intensity was detected after completion of each qPCR cycle and data was collected using the CFX Manager™ Software.

### VirCapSeq-VERT

For targeted viral detection, the capture technique VirCapSeq-VERT was performed as previously described (*83*). Low quality and poor complexity reads were filtered using PRINSEQ (v 0.20.2) and remaining reads were mapped with human host database using Bowtie 2 mapper 2.0.6 (*84*). Resulting reads were de novo assembled using MIRA (v4.0) assembler and homology search was performed on NCBI GenBank database (*85*). Both contigs and unique singletons from assembly were subjected to NCBI BLASTn, and unannotated sequences were further searched on non-redundant (*nr*) database using NCBI BLASTx. Based on BLASTn and BLASTx analysis viral sequences matching with Illumina reads and contigs, were downloaded from NCBI and used for mapping to recover partial or complete genomes.

### Viral RNA Sequencing

#### Wet lab methods for RNA and sequencing

RNA Libraries were made using TruSeq Stranded Total RNA with RiboErase (Illumina, USA) sample preparation. Illumina’s protocol was followed. Briefly, dependent on the extraction yield of RNA from CSF 1-100ng of RNA in 10ul were put into the prep, rRNA was depleted, cDNA and libraries were made and amplified for 15 cycles. The 130 libraries were pooled at 4nM, loaded for sequencing per the manufacturer’s protocol on the NovaSeq 6000 using the S2 2×100 flow cell.

#### PCR validation

Given the sensitivity limitations of both VirCapSeq and RNASeq for detection of virus, two qPCR assays were used to both confirm and screen for CMV in all of the preserved CSF and blood samples. For initial screening, a protocol previously described was used with a few optimizations (*86*). The reverse primer gB4 was modified to 5’-GGTGGTTGCCCAACAGGATT-3’ due to off target human amplification and Quantitect SybrGreen Mastermix (Qiagen, USA) was used following manufacturers protocol for 10 μL reaction volumes and 2 μL DNA input with PCR and cycling conditions recommended by (*87*). A standard curve was generated from 10 million to 1 million copies using a full length clone DNA of Homo cytomegalovirus gB (Sino Biologicals, Beijing, China) (*88, 89*). For confirmation of the initial screening all samples were ran using a TaqMan assay previously described (*90*). A standard curve was generated using a gBlock Gene Fragment of the UL54 region (IDT, Iowa, USA) from 10 million to 1 million copies. Technical replicates were evaluated in duplicate using each detection method. Samples were scored positive only if both replicates were positive in both assays.

PCR confirmation for human papilloma virus (HPV) was performed using the L1 Major Capsid Protein. HotStarTaq (Qiagen, Cat # 203203) was used in 25 µl reactions following manufacturer’s protocol with 0.5 µl of 10 µM primers and 5 µl of DNA. For Sanger Sequencing, a 1x AMpure bead p (Beckman Coulter Life Sciences) clean-up was performed eluting in 10 µl, mixed with forward and reverse primers then sent to Genewiz (NJ, USA). In the original PCR the product contained human sequences so nested PCR was performed as well using the same procedure in the first PCR reaction with a 1:10 dilution of the original PCR product. Sequencing was performed as well in the same manner. A positive HPV gBlock (IDT, USA) of the L1 major capsid protein gene diluted in human DNA was used as a positive control.

### Computational analysis and pipeline

We established an internal pipeline to detect human viral-specific reads within RNA-sequencing data. For quality pre-processing, reads and tails below a Phred score of 30 (*91, 92*), reads shorter than 75 bp, and residual adapter sequences are filtered using Trimmomatic (v. 0.36, 6) (*93*). Following pre-processing, unfiltered reads are aligned to hg38 with HISAT2 (v. 2.1.0) to eliminate any human-specific reads that may misalign to a manually curated virome reference database (*84*). Alignments are filtered by removing PCR duplicates, uniquely aligning reads (MAPQ score = 50), and removing tandem repeats with Picard (MarkDuplicates, v. 2.18.16, 8), Bedtools (v. 2.17.0, 10), Samtools, and Tandem Repeat Finder (v. 4.09, 9) to eliminate library generation bias and sequencing artifacts (Picard Tools, Broad Institute, http://broadinstitute.github.io/picard/) (*94, 95*). Finally, the number of virus-specific reads per viral strain within each sample are aggregated into a count matrix for downstream analysis.

## Supplementary Figures

**Supplementary Figure S1.**
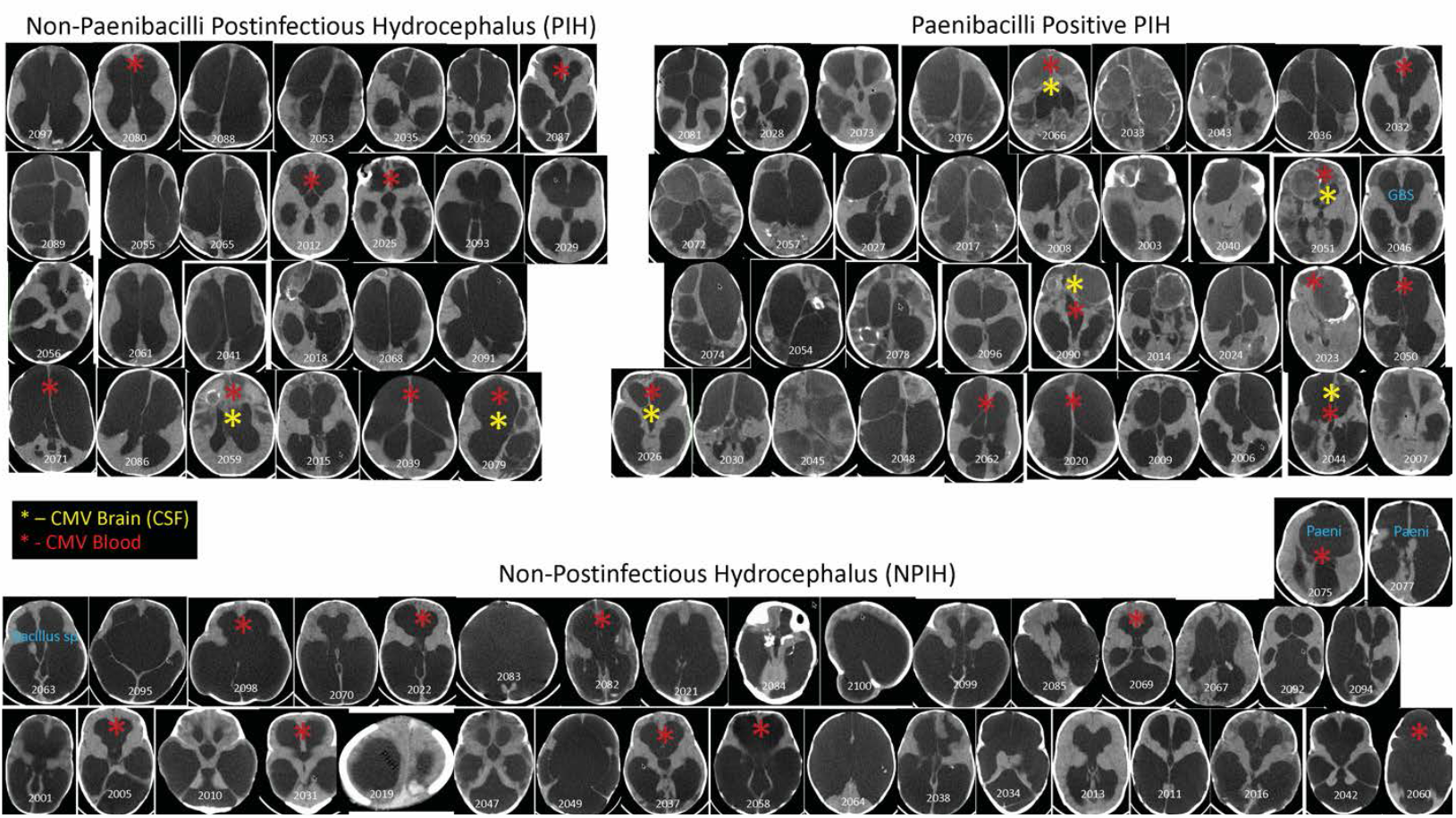
CT Scans. Composite of CT scans from all patients grouped by postinfectious hydrocephalus (PIH, upper groupings) and non-postinfectious (NPIH, lower group) hydrocephalus. The PIH group is further clustered by *Paenibacillus* negative (left) and positive (right). Yellow and red stars implicate if a patient was PCR positive for CMV in the CSF and blood respectively. Note that one PIH *Paenibacillus* positive patient (2002) has a missing CT scan. Two other patients met criteria for a bacterial identification for a putative pathogen, indicated as group B *Streptococcus* (GBS) in a case that was also *Paenibacillus* positive, and one case with *Bacillus sp*. in the NPIH group. Two patients in the NPIH group were positive for Paenibacilli (Paeni) using V1-V2 analysis. Note that no patient in the NPIH group that was CMV positive in the blood had CMV detected in the CSF.

**Supplementary Figure S2.**
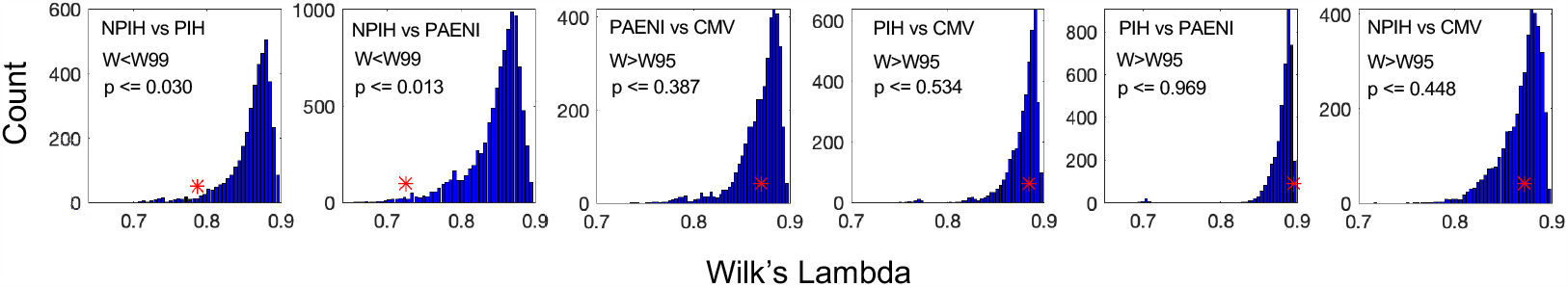
Spatial Statistics. Fisher’s canonical linear discrimination (LDA) contrasting the location of the groupings of cases shown in Fig. 1. Shown are Wilk’s lambda, W, reflecting the likelihood as the exponentiated sum of the log of the eigenvalues of the discriminant (*96*). W is chi-squared distributed, and values smaller (greater) than the 95th or 99th percentile confidence limits (W95 or W99 respectively) are statistically significant (nonsignificant). To bootstrap the significance of the result, 10,000 random permutations of the group assignments into PIH vs NPIH were performed, and W recalculated. The probability, p, that the W of the LDA on the original unpermuted data was significantly different (smaller) than the permuted values is shown. The W from the data is shown as a red asterisk (*) on each histogram.

**Supplementary Figure S3.**
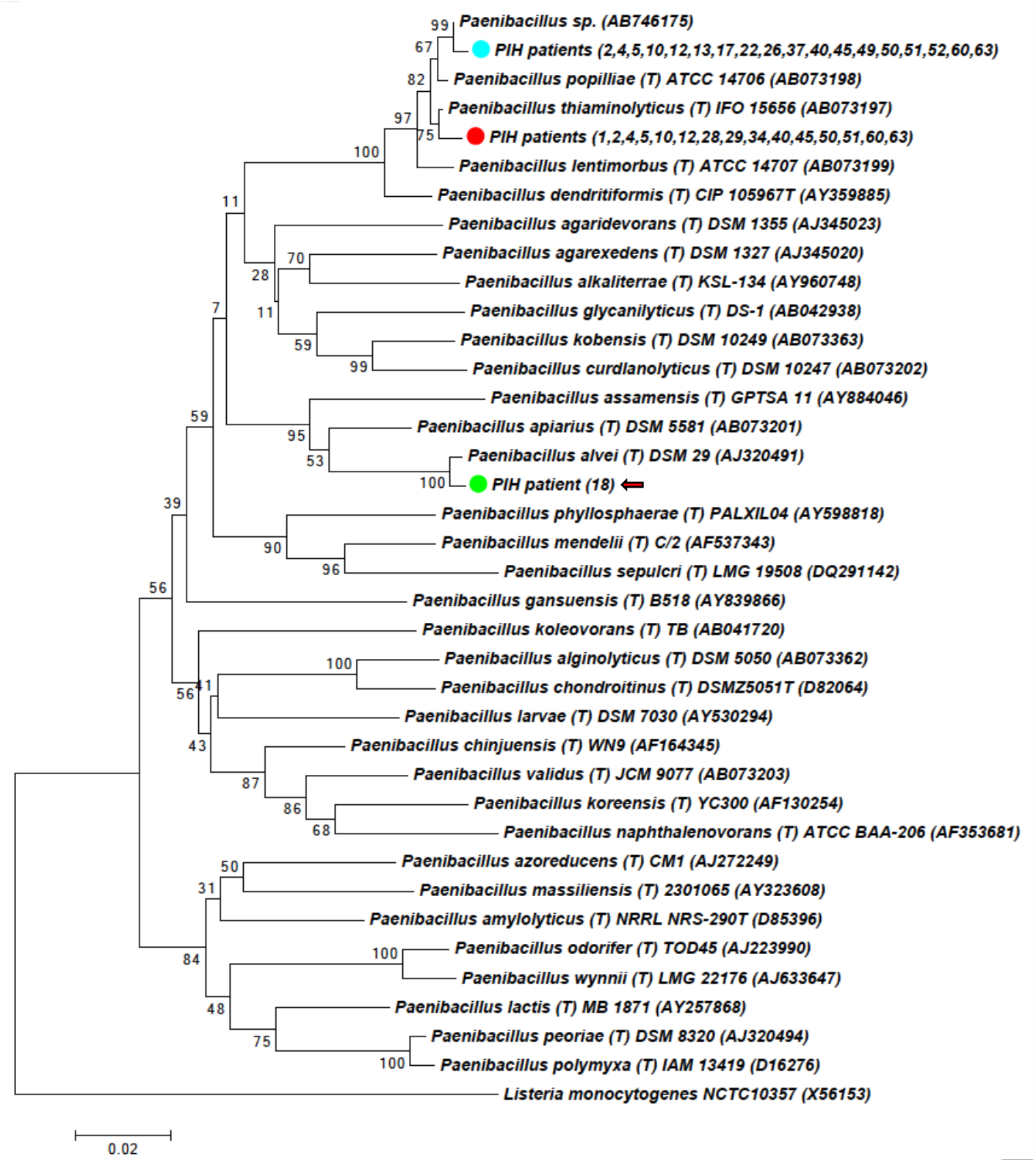
Phylogenetic tree for 16S V1-V4 sequences. Phylogenetic tree for 16S V1-V4 sequences obtained from CSF of PIH patients (colored circles) and related isolates. The optimal evolutionary history was inferred using the Neighbor-Joining method (*71*). The percentage of replicate trees in which the associated taxa clustered together in the bootstrap test (500 replicates) are shown next to the branches (*72*). The tree is drawn to scale, with branch lengths in the same units as those of the evolutionary distances used to infer the phylogenetic tree. The evolutionary distances were computed using the Kimura 2-parameter method (*73*), and are in the units of the number of base substitutions per site. The analysis involved 39 nucleotide sequences. All ambiguous positions were removed for each sequence pair. There were a total of 750 positions in the final dataset.

**Supplementary Figure S4.**
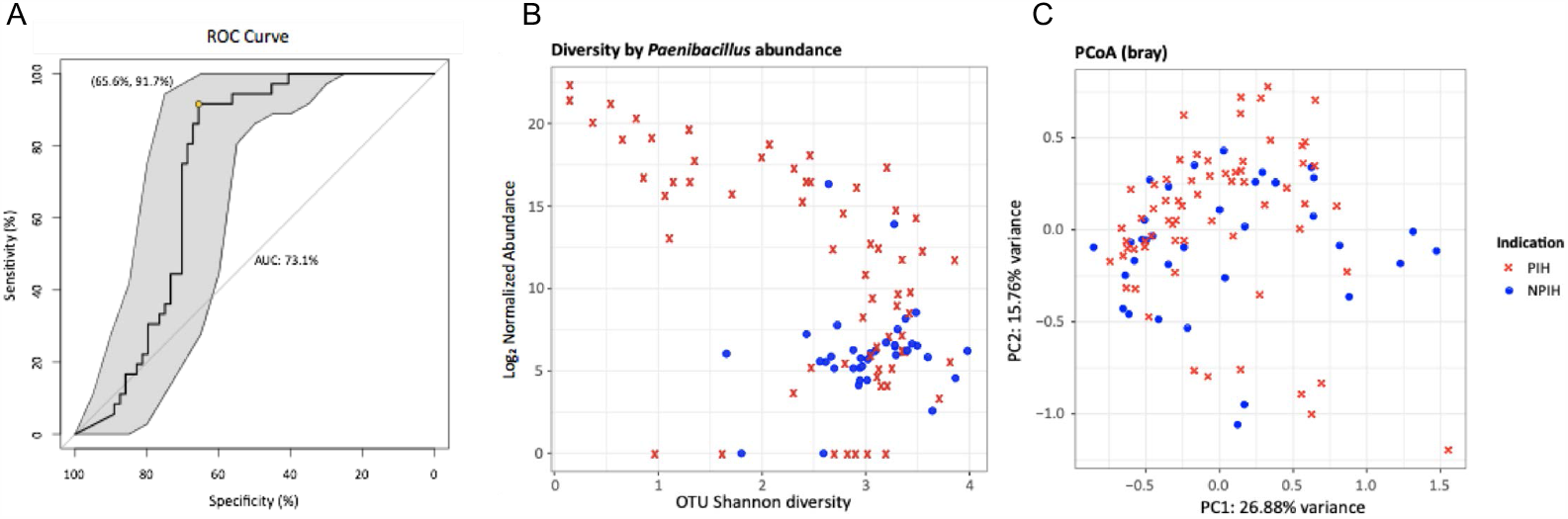
Further analysis of the microbial 16S community. Further analysis of the microbial 16S community. A) Receiver-operating-characteristic curve using *Paenibacillus* cumulative sum scaling (CSS) normalized abundance as the predictor for PIH or NPIH status. Area under the curve was 73.09% (95% DeLong CI = 63.19%-82.99%). B) Scatterplot of (y-axis) log2 normalized *Paenibacillus* 16S abundance by (x-axis) Shannon diversity estimates. C) Classical multidimensional scaling on the Bray-Curtis distance matrix of 16S abundances.

**Supplementary Figure S5.**
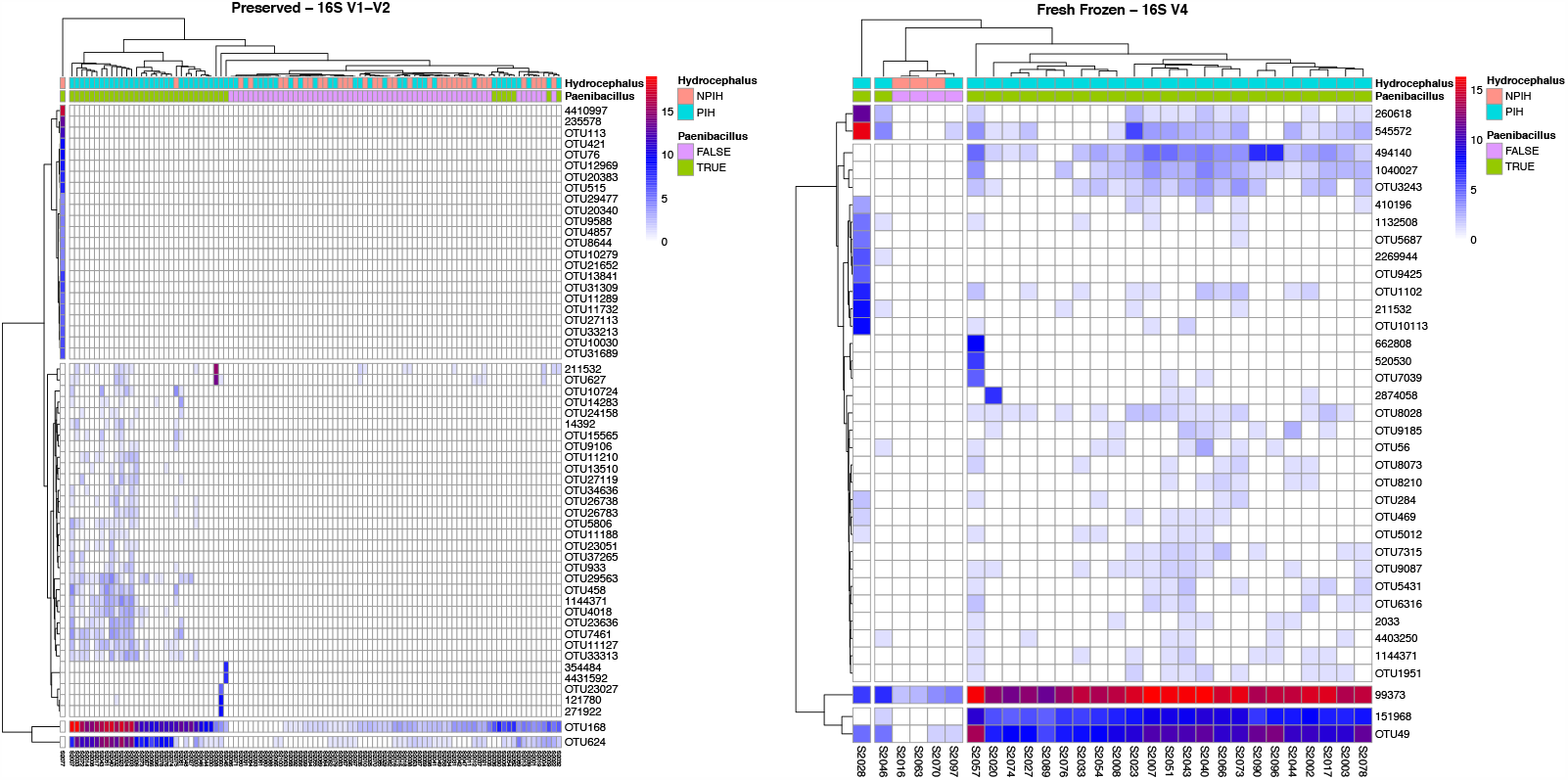
OTU Heatmaps. Log-transformed number of reads for each Operational Taxonomic Unit (OTU) annotated to the *Paenibacillus* genus. OTUs were subset to only those annotated to the *Paenibacillus* genus. The columns represent samples with hydrocephalus status (PIH/NPIH) and Paenibacillus positivity as defined by at least 50 reads within the respective dataset. Both the V1-V2 (left) and V4 (right) 16S sequencing have two dominant OTU sequence centers that are most represented among the positive *Paenibacillus* (green) patients with the exception of an NPIH positive case that clusters separately in the V1-V2 sequencing cohort. These OTU centers have high sequence similarity to the *P. thiaminolyticus* Mbale clinical isolate while not having sequence similarity ≥ 97% to the other two *Paenibacillus* isolates. Specifically, the two most abundant OTU sequence centers in V4, 99373 and 151968, align at 100% and 99.6% respectively along with three other less abundant but present centers OTUs 545572, 1144371, and 494140 that align at >99%. For V1-V2, OTU 168 and 624 dominate the signal and align to the clinical isolate at 100% and 97% respectively.

**Supplementary Figure S6.**
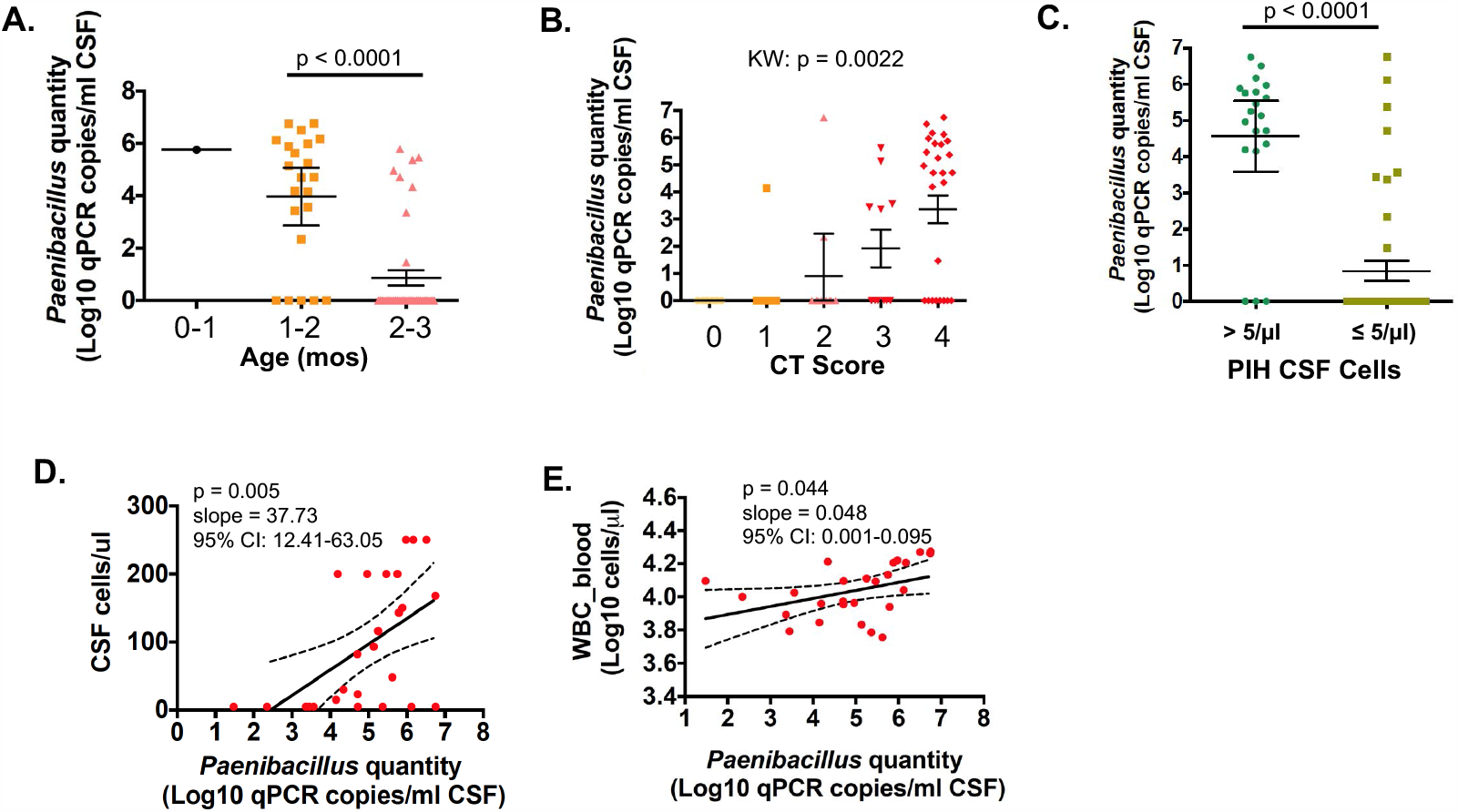
qPCR vs Age, CT Score, and Cell Counts. Distribution of *Paenibacillus* quantity as determined by qPCR in CSF by age (**A**) and CT score (**B**). Quantity of *Paenibacillus* in CSF from PIH patients with >5 cells/µl CSF compared to PIH patients with ≤ 5 cells/µl CSF (**C**). Relationship between *Paenibacillus* quantity in CSF and CSF cell counts (cells/µl) in PIH patients with quantifiable *Paenibacillus* by qPCR (**D**). Relationship between *Paenibacillus* quantity in CSF and WBC counts in blood (cells/µl) in PIH patients with quantifiable *Paenibacillus* by qPCR (**E**). The linear regression line estimated by a univariate model for CSF cells (**D**) or WBC counts (**E**) with *Paenibacillus* quantity as the primary covariate is shown (solid line in **D, E**). p-values shown were derived from the Mann-Whitney U test (**A, C**), the Kruskal-Wallace test (**B**) and univariate linear regression (**D, E**). The estimated increase in CSF cells or WBC counts for one log10 fold increase in Paenibacillus quantity and associated 95% CI is indicated (slope and 95% CI, **D** and **E**).

**Supplementary Figure S7.**
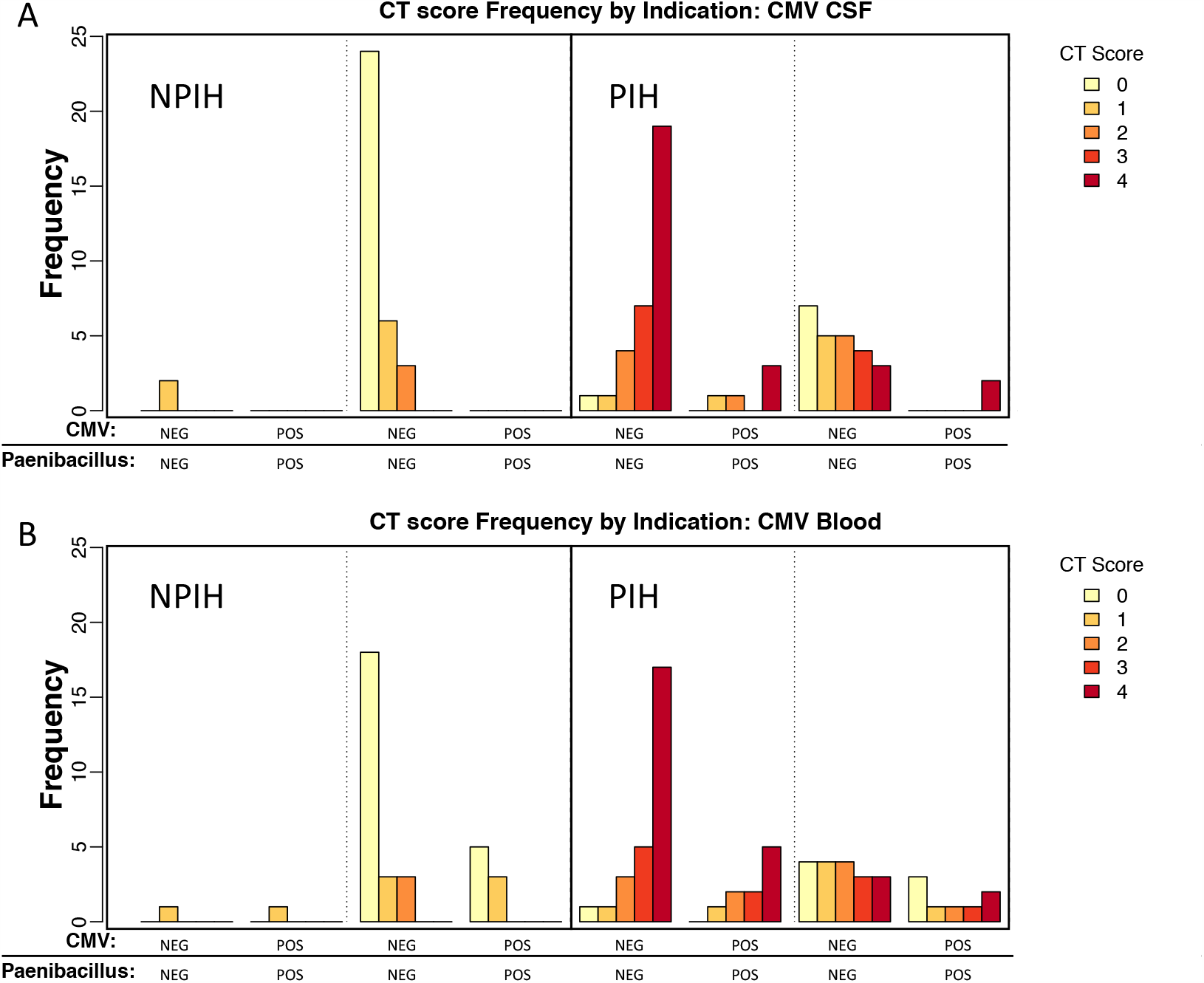
CT Score Frequencies. A) CT scores as a function of CMV infection status, positive or negative, in CSF. CT score status further stratified by clinical indication, PIH or NPIH, and whether CSF 16S sequencing was positive for *P. thiaminolyticus*, and whether CMV was positive or negative in CSF. B) CT scores as a function of CMV infection status, positive or negative, in blood. CT score status further stratified by clinical indication, PIH or NPIH, and whether CSF 16S sequencing was positive for *P. thiaminolyticus*, and whether CMV was positive or negative in blood.

## Supplementary Tables

**Supplementary Table S1.**
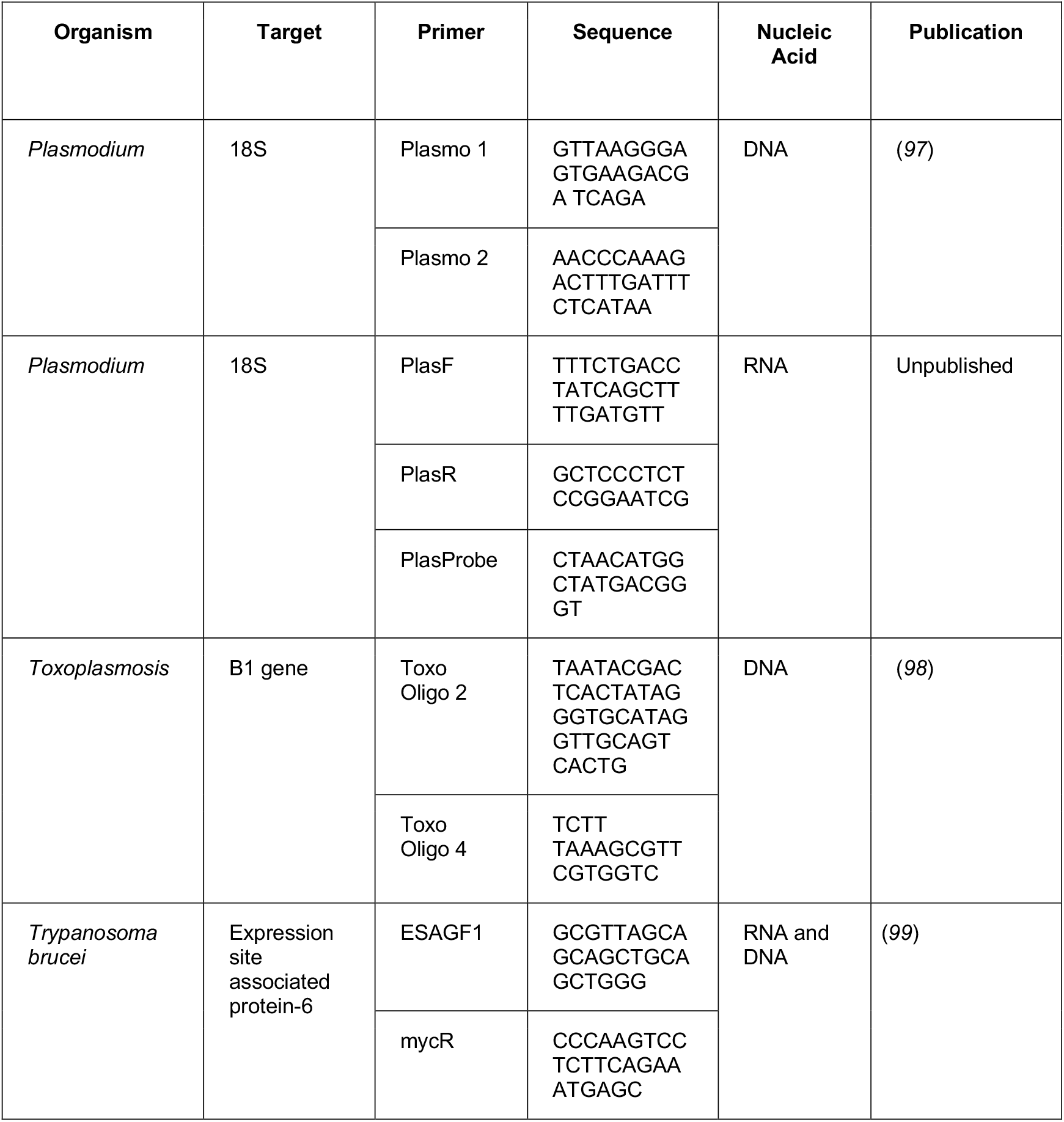
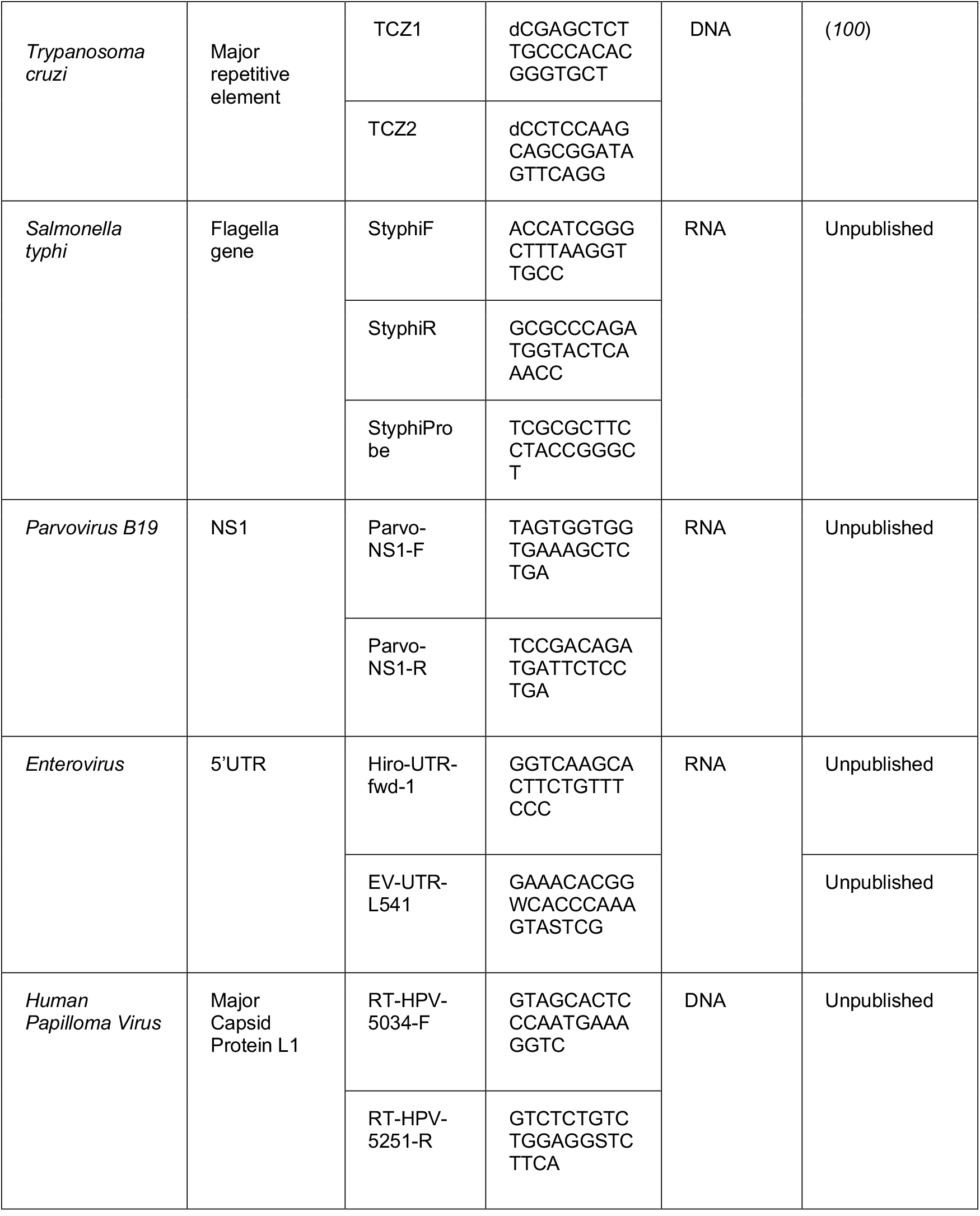
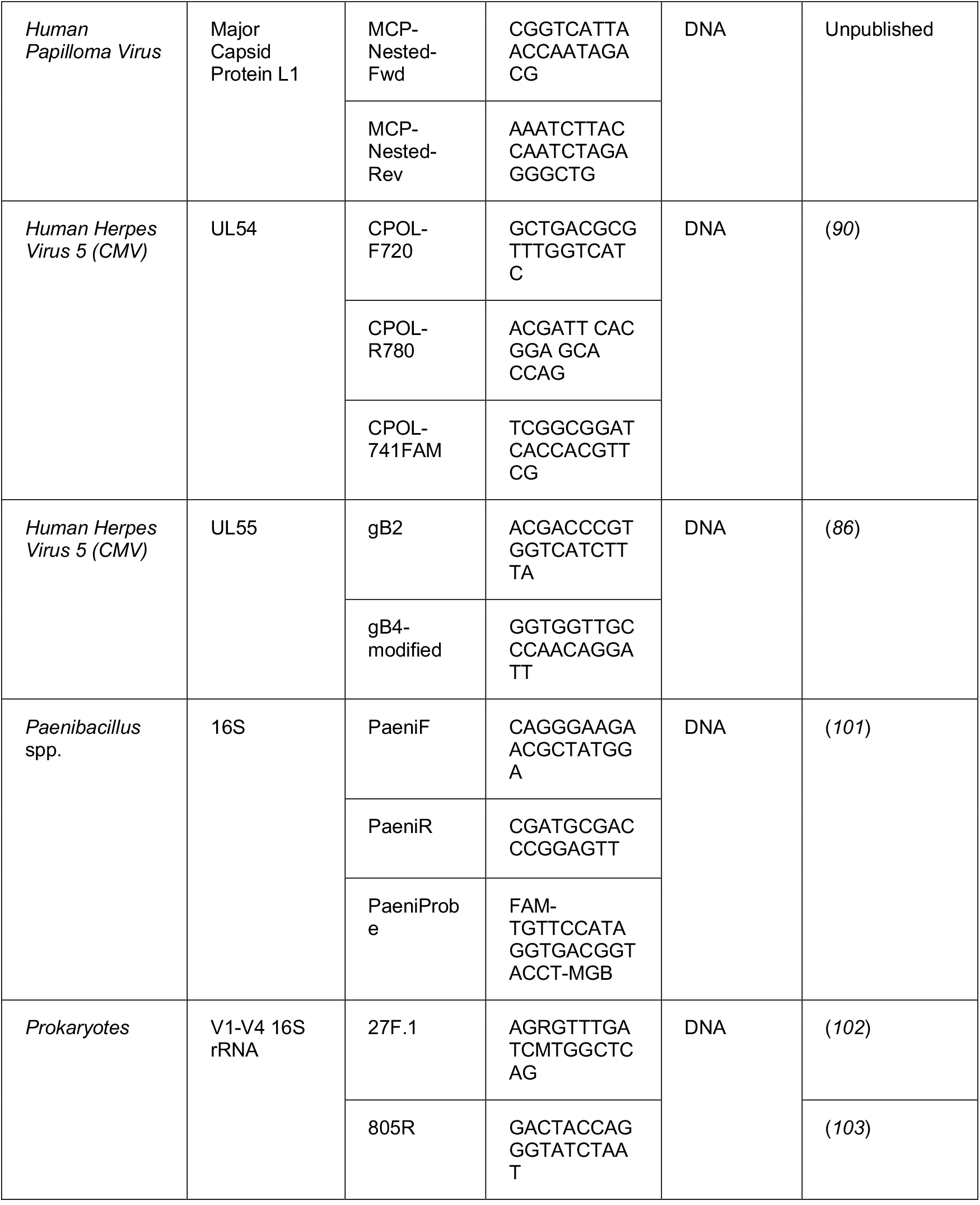
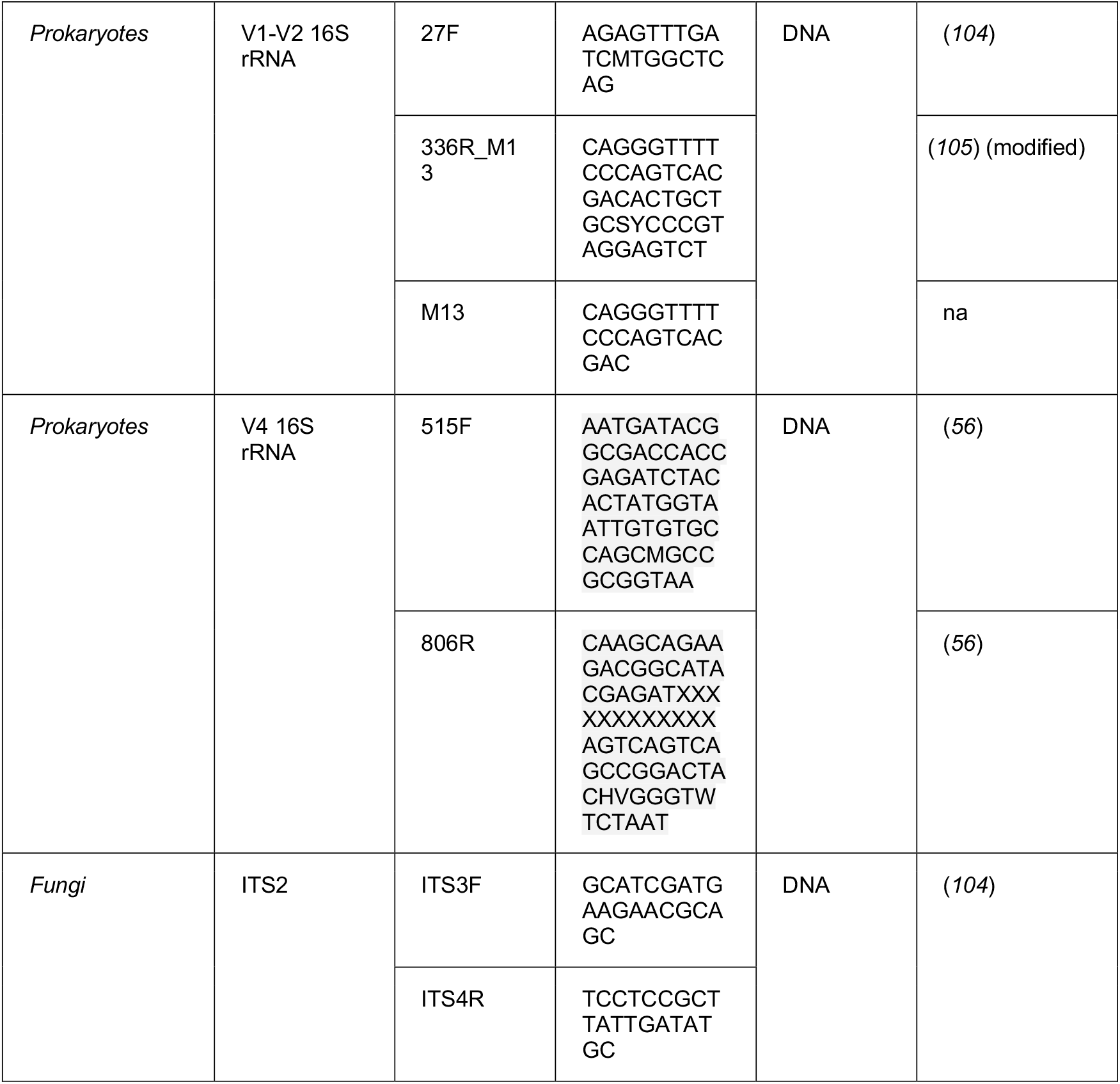
Primer Table. Primer table for all PCR to identify microbiological DNA or RNA present in the CSF or Blood.

**Supplementary Table S2.** Fresh Frozen PCR and Sequencing. Summary of results based on conventional PCR (cPCR) using 16S V1-V4 primers with cloning and sequencing, *Paenibacillus* genus-specific qPCR, and V4 MiSeq on DNA obtained from fresh frozen CSF from each PIH (n=64) and NPIH (n=36) subject and concordance between the three assays for the presence of *Paenibacillus*.

| Sample Number | Group | cPCR 16S V1-V4 amplification | cPCR 16S V1-V4 (Cloning/Sequencing Results) | *Paenibacillus* genus-specific qPCR (16S copies/mL CSF) | V4 MiSeq (Dominant taxa) | Assay Concordance (# Assays demonstrating *Paenibacillus*) |
| --- | --- | --- | --- | --- | --- | --- |
| 2002 | PIH | Positive | *Paenibacillus thiaminolyticus* or *popilliae* | 2.33E+05 | *Paenibacillus* sp. | 3 |
| 2003 | PIH | Positive | *Paenibacillus thiaminolyticus* or *popilliae* | 5.07E+04 | *Paenibacillus* sp. | 3 |
| 2006 | PIH | Negative | no amplification | 0 | no amplification | 0 |
| 2007 | PIH | Positive | *Paenibacillus thiaminolyticus* or *popilliae* | 5.59E+06 | *Paenibacillus* sp. | 3 |
| 2008 | PIH | Positive | *Paenibacillus thiaminolyticus* or *popilliae* | 1.79E+05 | *Paenibacillus* sp. | 3 |
| 2009 | PIH | Negative | no amplification | 0 | no amplification | 0 |
| 2012 | PIH | Negative | no amplification | 0 | no amplification | 0 |
| 2014 | PIH | Negative | no amplification | 0 | no amplification | 0 |
| 2015 | PIH | Negative | no amplification | 0 | no amplification | 0 |
| 2017 | PIH | Positive | *Paenibacillus thiaminolyticus* or *popilliae* | 1.49E+06 | *Paenibacillus* sp. | 3 |
| 2018 | PIH | Negative | no amplification | 0 | no amplification | 0 |
| 2020 | PIH | Positive | *Paenibacillus thiaminolyticus* or *popilliae* | 3.63E+03 | *Paenibacillus* sp. | 3 |
| 2023 | PIH | Positive | *Paenibacillus thiaminolyticus* or *popilliae* | 3.23E+06 | *Paenibacillus* sp. | 3 |
| 2024 | PIH | Negative | no amplification | 0 | no amplification | 0 |
| 2025 | PIH | Negative | no amplification | 0 | no amplification | 0 |
| 2026 | PIH | Negative | no amplification | 1.41E+04 | no amplification | 1 |
| 2027 | PIH | Positive | *Paenibacillus thiaminolyticus* or *popilliae* | 5.16E+04 | *Paenibacillus* sp. | 3 |
| 2028 | PIH | Positive | *Paenibacillus alvei* | 0* | *Paenibacillus alvei* | 2 |
| 2029 | PIH | Negative | no amplification | 0 | no amplification | 0 |
| 2030 | PIH | Negative | no amplification | 0 | no amplification | 0 |
| 2032 | PIH | Negative | no amplification | 0 | no amplification | 0 |
| 2033 | PIH | Positive | *Paenibacillus thiaminolyticus* or *popilliae* | 5.67E+05 | *Paenibacillus* sp. | 3 |
| 2035 | PIH | Negative | no amplification | 0 | no amplification | 0 |
| 2036 | PIH | Negative | no amplification | 0 | no amplification | 0 |
| 2039 | PIH | Negative | no amplification | 0 | no amplification | 0 |
| 2040 | PIH | Positive | *Paenibacillus thiaminolyticus* or *popilliae* | 4.23E+05 | *Paenibacillus* sp. | 3 |
| 2041 | PIH | Negative | no amplification | 0 | no amplification | 0 |
| 2043 | PIH | Positive | *Paenibacillus thiaminolyticus* or *popilliae* | 5.68E+06 | *Paenibacillus* sp. | 3 |
| 2044 | PIH | Positive | *Paenibacillus thiaminolyticus* or *popilliae* | 2.20E+02 | *Paenibacillus* sp. | 3 |
| 2045 | PIH | Negative | no amplification | 0 | no amplification | 0 |
| 2046 | PIH | Positive | *Streptococcus agalactiae* | 0 | *Streptococcus* sp. | 0 |
| 2048 | PIH | Negative | no amplification | 2.91E+01 | no amplification | 1 |
| 2050 | PIH | Positive | *Alkanindiges* sp. | 0 | no amplification | 0 |
| 2051 | PIH | Positive | *Paenibacillus thiaminolyticus* or *popilliae* | 1.33E+06 | *Paenibacillus* sp. | 3 |
| 2052 | PIH | Negative | no amplification | 0 | no amplification | 0 |
| 2053 | PIH | Negative | no amplification | 0 | no amplification | 0 |
| 2054 | PIH | Positive | *Paenibacillus thiaminolyticus* or *popilliae* | 5.22E+04 | *Paenibacillus* sp. | 3 |
| 2055 | PIH | Negative | no amplification | 0 | no amplification | 0 |
| 2056 | PIH | Negative | no amplification | 0 | no amplification | 0 |
| 2057 | PIH | Positive | *Paenibacillus thiaminolyticus* or *popilliae* | 2.22E+04 | *Paenibacillus* sp. | 3 |
| 2059 | PIH | Negative | no amplification | 0 | no amplification | 0 |
| 2061 | PIH | Negative | no amplification | 0 | no amplification | 0 |
| 2062 | PIH | Negative | no amplification | 0 | no amplification | 0 |
| 2065 | PIH | Negative | no amplification | 0 | no amplification | 0 |
| 2066 | PIH | Positive | *Paenibacillus thiaminolyticus* or *popilliae* | 9.22E+04 | *Paenibacillus* sp. | 3 |
| 2068 | PIH | Negative | no amplification | 0 | no amplification | 0 |
| 2071 | PIH | Negative | no amplification | 0 | no amplification | 0 |
| 2072 | PIH | Negative | no amplification | 9.46E+05 | no amplification | 1 |
| 2073 | PIH | Positive | *Paenibacillus thiaminolyticus* or *popilliae* | 1.37E+05 | *Bacteroides* and *Paenibacillus* sp. | 3 |
| 2074 | PIH | Positive | *Paenibacillus thiaminolyticus* or *popilliae* | 6.19E+05 | *Paenibacillus* sp. | 3 |
| 2076 | PIH | Positive | *Paenibacillus thiaminolyticus* or *popilliae* | 1.56E+04 | *Paenibacillus* sp. | 3 |
| 2078 | PIH | Positive | *Paenibacillus thiaminolyticus* or *popilliae* | 7.64E+05 | *Paenibacillus* sp. | 3 |
| 2079 | PIH | Negative | no amplification | 0 | no amplification | 0 |
| 2080 | PIH | Negative | no amplification | 0 | no amplification | 0 |
| 2081 | PIH | Negative | no amplification | 0 | no amplification | 0 |
| 2086 | PIH | Negative | no amplification | 0 | no amplification | 0 |
| 2087 | PIH | Negative | no amplification | 0 | no amplification | 0 |
| 2088 | PIH | Negative | no amplification | 0 | no amplification | 0 |
| 2089 | PIH | Positive | Mixed taxa | 2.33E+03 | *Staphylococcus* and *Paenibacillus* sp. | 2 |
| 2090 | PIH | Positive | *Paenibacillus thiaminolyticus* or *popilliae* | 2.93E+05 | *Paenibacillus* sp. | 3 |
| 2091 | PIH | Negative | no amplification | 0 | no amplification | 0 |
| 2093 | PIH | Negative | no amplification | 0 | no amplification | 0 |
| 2096 | PIH | Positive | *Paenibacillus thiaminolyticus* or *popilliae* | 2.79E+03 | *Paenibacillus* sp. | 3 |
| 2097 | PIH | Positive | Mixed taxa | 0 | *Streptococcus* sp. | 0 |
| 2001 | NPIH | Negative | no amplification | 0 | no amplification | 0 |
| 2004 | NPIH | Negative | no amplification | 0 | no amplification | 0 |
| 2005 | NPIH | Negative | no amplification | 0 | no amplification | 0 |
| 2010 | NPIH | Negative | no amplification | 0 | no amplification | 0 |
| 2011 | NPIH | Negative | no amplification | 0 | no amplification | 0 |
| 2013 | NPIH | Negative | no amplification | 0 | no amplification | 0 |
| 2016 | NPIH | Positive | Clostridiales sp. | 0 | *Burkholderia* sp. | 0 |
| 2019 | NPIH | Negative | no amplification | 0 | no amplification | 0 |
| 2021 | NPIH | Negative | no amplification | 0 | no amplification | 0 |
| 2022 | NPIH | Negative | no amplification | 0 | no amplification | 0 |
| 2031 | NPIH | Negative | no amplification | 0 | no amplification | 0 |
| 2034 | NPIH | Negative | no amplification | 0 | no amplification | 0 |
| 2037 | NPIH | Negative | no amplification | 0 | no amplification | 0 |
| 2038 | NPIH | Negative | no amplification | 0 | no amplification | 0 |
| 2042 | NPIH | Negative | no amplification | 0 | no amplification | 0 |
| 2047 | NPIH | Negative | no amplification | 0 | no amplification | 0 |
| 2049 | NPIH | Negative | no amplification | 0 | no amplification | 0 |
| 2058 | NPIH | Negative | no amplification | 0 | no amplification | 0 |
| 2060 | NPIH | Negative | no amplification | 0 | no amplification | 0 |
| 2063 | NPIH | Positive | *Bacillus* sp. | 0 | *Bacillus* sp. | 0 |
| 2064 | NPIH | Negative | no amplification | 0 | no amplification | 0 |
| 2067 | NPIH | Negative | no amplification | 0 | no amplification | 0 |
| 2069 | NPIH | Negative | no amplification | 0 | no amplification | 0 |
| 2070 | NPIH | Positive | Mixed taxa | 0 | Clostridiales sp. | 0 |
| 2075 | NPIH | Negative | no amplification | 0 | no amplification | 0 |
| 2077 | NPIH | Negative | no amplification | 0 | no amplification | 0 |
| 2082 | NPIH | Negative | no amplification | 0 | no amplification | 0 |
| 2083 | NPIH | Negative | no amplification | 0 | no amplification | 0 |
| 2084 | NPIH | Negative | no amplification | 0 | no amplification | 0 |
| 2085 | NPIH | Negative | no amplification | 0 | no amplification | 0 |
| 2092 | NPIH | Negative | no amplification | 0 | no amplification | 0 |
| 2094 | NPIH | Negative | no amplification | 0 | no amplification | 0 |
| 2095 | NPIH | Negative | no amplification | 0 | no amplification | 0 |
| 2098 | NPIH | Negative | no amplification | 0 | no amplification | 0 |
| 2099 | NPIH | Negative | no amplification | 0 | no amplification | 0 |
| 2100 | NPIH | Negative | no amplification | 0 | no amplification | 0 |
| * Note, genus-specific qPCR primers for *Paenibacillus* would not be expected to amplify *Paenibacillus alvei* due to substantial mismatches in the primer binding sites of this species |  |  |  |  |  |  |

**Supplementary Table S3.** Differential abundance. Statistical results from 16S Differential Abundance Testing using V1-V2. Counts were aggregated to the genera level and a log-normal regression model was applied using limma (*106*) with abundance as the outcome variable and indication status as the independent variable. The ten genera with the highest B-statistic are reported.

| **Taxa** | **logFC** | **AveExp** | **t** | **p.value** | **adjusted p.value** | **B** |
| --- | --- | --- | --- | --- | --- | --- |
| g__Paenibacillus | 4.8292 | 8.0664 | 4.1496 | 0.0001 | 0.0111 | 1.5032 |
| g__Tepidimonas | 3.0803 | 8.4413 | 3.4537 | 0.0008 | 0.0645 | -0.6682 |
| g__Rothia | 2.1504 | 1.9001 | 2.9372 | 0.0041 | 0.2178 | -2.0791 |
| g__Micrococcus | 1.6369 | 1.6634 | 2.4435 | 0.0163 | 0.6476 | -3.2479 |
| g__Anaerococcus | -1.2173 | 1.1405 | -2.1411 | 0.0347 | 0.6806 | -3.8703 |
| g__Anoxybacillus | 1.0416 | 0.7633 | 2.0259 | 0.0454 | 0.6806 | -4.088 |
| g__Moraxella | -0.4952 | 0.1783 | -1.914 | 0.0585 | 0.6806 | -4.2887 |
| g__Actinomycetospora | -0.4519 | 0.1627 | -1.8299 | 0.0702 | 0.6806 | -4.4328 |
| g__Gordonia | 0.4937 | 0.4333 | 1.8238 | 0.0712 | 0.6806 | -4.4428 |
| no_match | -0.6577 | 12.7805 | -1.8053 | 0.074 | 0.6806 | -4.4736 |

**Supplementary Table S4.** Viral Results. Summary of viral analysis using VirCapSeq, PCR, and RNAseq. All 100 CSF samples were tested with VirCapSeq-VERT and RNASeq. VirCapSeq-VERT detected evidence of 12 viruses present across 36 samples and RNASeq detected Humans Herpesvirus 5 (CMV) in 4 samples. The only virus that was confirmed through PCR at high abundance in our samples was CMV, for which we confirmed 5 of the 8 VirCapSeq-VERT samples and 3 of the 4 RNASeq positive CSF samples. CMV was only identified in the CSF from CMV blood positive PIH cases. PCR testing was performed for CMV and HPV.

| Viral Summary Results | VirCapSeq-VERT |  | RNA-Seq |  | PCR |  |  |  |
| --- | --- | --- | --- | --- | --- | --- | --- | --- |
|  | PIH (64) | NPIH (36) | PIH (64) | NPIH (36) | PIH (n) |  | NPIH (n) |  |
| Virus | CSF |  | CSF |  | Blood | CSF | Blood | CSF |
| Human Herpesvirus 5 (CMV) | 8 | 0 | 4 | 0 | 18 (64) | 8 (64) | 9 (35) | 0 (36) |
| Human Papillomavirus | 8 | 8 | 0 | 0 | 0 (11) | 0 (11) | 0 (8) | 0 (8) |
| Human Pegivirus / GBV-C | 3 | 1 | 0 | 0 |  |  |  |  |
| Human Herpesvirus 6A | 1 | 0 | 0 | 0 |  |  |  |  |
| Human Herpesvirus 6B | 1 | 2 | 0 | 0 |  |  |  |  |
| Human Rotavirus | 1 | 1 | 0 | 0 |  |  |  |  |
| Torque Teno Virus | 2 | 1 | 0 | 0 |  |  |  |  |
| Human Parvovirus (B19/V9) | 1 | 0 | 0 | 0 |  |  |  |  |
| Norovirus | 1 | 0 | 0 | 0 |  |  |  |  |
| Merkel Cell Polyomavirus | 0 | 1 | 0 | 0 |  |  |  |  |
| Circovirus | 0 | 1 | 0 | 0 |  |  |  |  |
| No Viral Reads | 43 | 21 | 60 | 36 |  |  |  |  |

**Supplementary Table S5.** Demographics with and without Paenibacillus. Comparison of demographic and clinical attributes between three groups: NPIH, PIH patients infected with *Paenibacillus* and PIH patients without *Paenibacillus*. PIH patients with *Paenibacillus* were more likely to be younger compared to those without *Paenibacillus* but there were no gender differences between these three groups. To investigate if the biomarkers of active inflammation were different between the three groups, we analyzed white blood cell (WBC) count. WBC from peripheral blood and CSF were significantly higher in the PIH group with *Paenibacillus* compared to other groups. However, the *Paenibacillus* group did not show a difference in hemoglobin or hematocrit concentration when compared to PIH without *Paenibacillus*. Preoperative computed tomography (CT) scans were available for 98 patients (98%). Of these, patients in the PIH group with *Paenibacillus* were more likely to have a higher CT scan score indicative of brain abscess, calcifications, loculations and septations (p<0.0001). However, there were no significant differences in gender and human immunodeficiency virus (HIV) frequency between the groups. Continuous demographic variables were evaluated using the non-parametric Wilcoxon rank-sum (2-group comparisons) and Kruskal-Wallis (>2 groups) tests following Shapiro-Wilk’s test for normality, unless otherwise stated. Fisher’s exact test was performed for categorical variables.

| Characteristics | All patients<br>n=100 | PIH with<br>*Paenibacillus*<br>n=38 | PIH without<br>*Paenibacillus*<br>n=26 | NPIH<br>n=36 |
| --- | --- | --- | --- | --- |
| Age in days, mean (SD) | 57 (24) | 59 (17) | 75 (11) | 43 (27) |
| Sex |  |  |  |  |
| Male (%) | 51 (51) | 21 (55) | 14 (54) | 16 (44) |
| Female (%) | 49 (49) | 17 (45) | 12 (46) | 20 (56) |
| Peripheral blood WBC<br>[1.0 x 10<sup>3</sup>]/μL, mean (SD) | 10.2 (3.6) | 12 (4) | 9.8 (3) | 8.8 (2.6) |
| Lymphocytes % |  | 56.8 (12.1) | 65.4 (8.8) | 59.3 (12.0) |
| Granulocytes % (SD) |  | 33.6 (13.6) | 26.0 (8.4) | 31.1 (10.9) |
| CSF WBC/μL, mean (SD)‡ | 30 (62) | 71 (87) | 5 (0) | 5 (0.5) |
| Glucose mg/dL (SD) |  | 32.4 (16.8) | 30.5 (7.6) | 38.8 (13.5) |
| Protein mg/dL (SD) |  | 166.2 (108.3) | 70.4 (60.4) | 37.7 (33.1) |
| Hemoglobin g/dl, mean (SD) | 11.5 (2.2) | 10.8 (1.3) | 10.5 (1.3) | 13.0 (2.8) |
| Hematocrit %, mean (SD) | 36.8 (7.4) | 34.4 (4.1) | 33.6 (4.2) | 41.6 (9.3) |
| CT scan scoring*<br>[no. and % in each category] |  |  |  |  |
| 0 | 32 (33) | 1 (3) | 7 (27) | 24 (69) |
| 1 | 15 (15) | 2 (5) | 5 (19) | 8 (23) |
| 2 | 13 (13) | 5 (14) | 5 (19) | 3 (9) |
| 3 | 11 (11) | 7 (19) | 4 (16) | 0 (0) |
| 4 | 27 (26) | 22 (59) | 5 (19) | 0 (0) |
| HIV Exposure status [no. and %] |  |  |  |  |
| Yes | 5 (5) | 1 (3) | 2 (8) | 2 (6) |
| No | 95 (95) | 37 (97) | 24 (92) | 34 (94) |

\* n=98
‡ n=99

**Supplementary Table S6.**
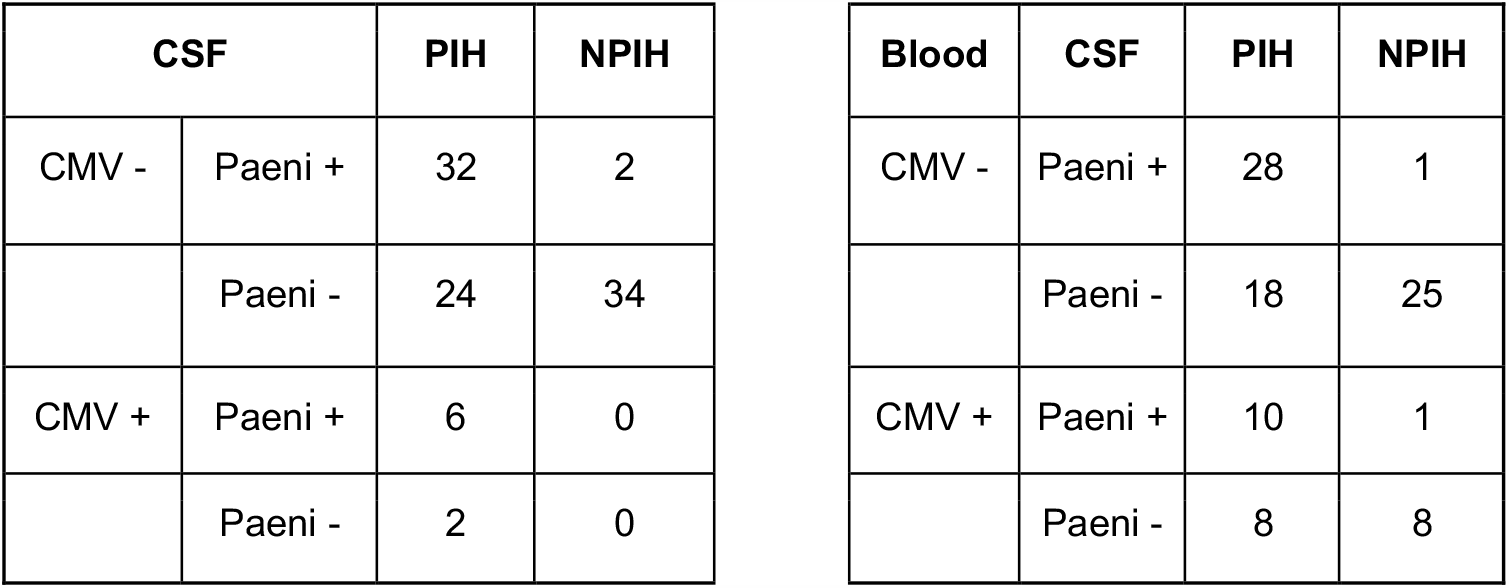
Paenibacillus vs CMV. Contingency tables for the prevalence of *Paenibacillus* positivity defined by 16S and the detection of CMV found in CSF or blood respectively. Interestingly, we did not observe any CMV positive CSF in NPIH samples.

**Supplementary Table S7.** CT Score vs CMV. Table of CT score composite features cross tabulated with CMV CSF status. Within the CMV negative population, 26%, 48%, 53%, and 47% had signs of abscesses, calcifications, loculations and debris respectively. In contrast the CMV positive population, albeit with small numbers, 71% and 86% of patients had signs of abscesses or calcifications, loculations, or debris respectively. Controlling for Paenibacillus presence in an ordinal logistic regression did not demonstrate a significant odds ratio relating CMV positivity to elevated CT scan scores OR (95% CI) = 3.30 (0.52, 29.66).

| **Loculations\CMV CSF** | **Negative** | **Positive** | **Totals** |
| --- | --- | --- | --- |
| Negative | 43 | 1 | 44 |
| Positive | 48 | 6 | 54 |
| Totals | 91 | 7 | 98 |

| **Debris\CMV CSF** | **Negative** | **Positive** | **Totals** |
| --- | --- | --- | --- |
| Negative | 48 | 1 | 49 |
| Positive | 43 | 6 | 49 |
| Totals | 91 | 7 | 98 |

| **Calcification\CMV CSF** | **Negative** | **Positive** | **Totals** |
| --- | --- | --- | --- |
| Negative | 47 | 1 | 48 |
| Positive | 44 | 6 | 50 |
| Totals | 91 | 7 | 98 |

| **Abscess\CMV CSF** | **Negative** | **Positive** | **Totals** |
| --- | --- | --- | --- |
| Negative | 67 | 2 | 69 |
| Positive | 24 | 5 | 29 |
| Totals | 91 | 7 | 98 |

**Supplementary Table S8.** Culture Media. Detailed description of the six different culture media with the respective inoculum volume, conditions and duration of culture. BBL™ Chocolate Agar (CHOC), BBL™ Sheep blood Agar (BAP), BBL™ MacConkey Agar (MAC), Chopped Meat Glucose Broth (CMG), BD BACTEC™ Peds Plus™ medium (PEDS), BD BACTEC™ Lytic Anaerobic medium (ANA).

| **Media** | **Volume of inoculum (µl)** | **Conditions** | **Duration (Days)** |
| --- | --- | --- | --- |
| **CHOC** | 100 | 37°C, 5% CO<sub>2</sub> | 7 |
| **BAP** | 100 | 37°C, 5% CO<sub>2</sub> | 7 |
| **MAC** | 100 | 37°C, 5% CO<sub>2</sub> | 7 |
| **CMG** | 200 | 37°C, Ambient air | 14 |
| **PEDS** | 250 | 37°C, BACTEC instrument | 14 |
| **ANA** | 50 | 37°C, BACTEC instrument | 14 |

**Supplementary Table S9.**
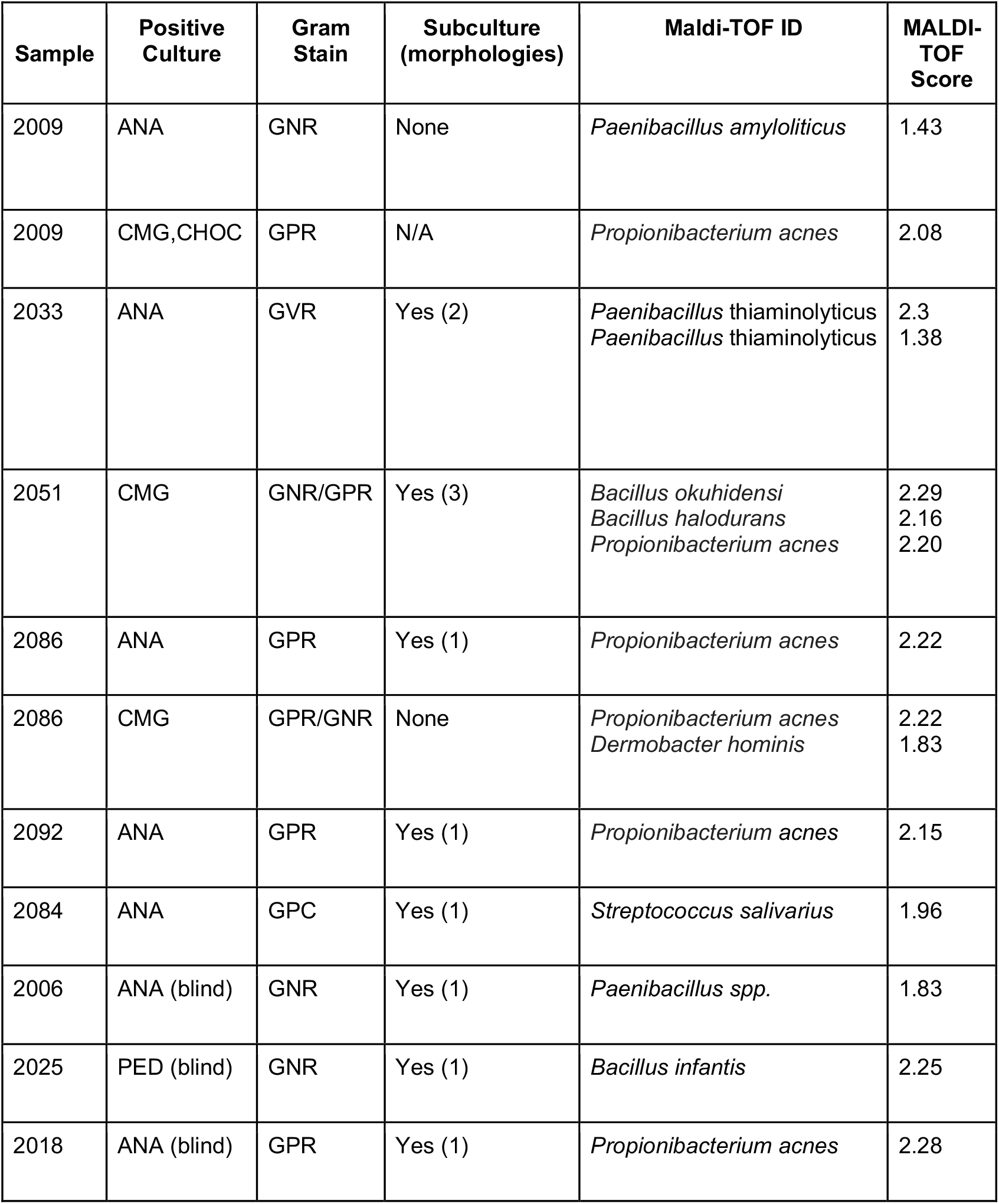
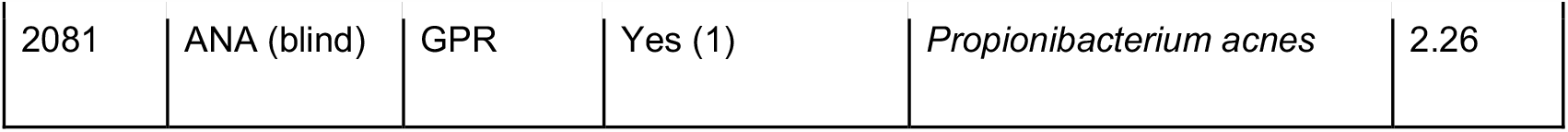
Culture Results. The culture and subculture results from 100 fresh frozen CSF patient samples. All 100 fresh frozen CSF samples were inoculated in 6 different media outlined in Table S8 and subcultured. Successful subcultures and solid media cultures were tested with MALDI-TOF for identification using 1.7 and 2.0 cut offs for genus and species identification respectively. For samples that were unable to grow on solid media MALDI-TOF was performed on the liquid culture. Further, negative liquid cultures were blindly subcultured (blind) and positive cultures were also tested with MALDI-TOF. (Supplementary Methods: Characterization for Organisms Recovered from Culture).Gram negative rods (GNR), Gram positive rods (GPR), Gram variable rods (BVR), Gram positive cocci (GPC), BBLTM Chocolate Agar (CHOC), BBLTM Sheep blood Agar (BAP), BBLTM MacConkey Agar (MAC), Chopped Meat Glucose Broth (CMG), BD BACTEC™ Peds Plus™ medium (PEDS), BD BACTEC™ Lytic Anaerobic medium (ANA).

**Supplementary Table S10.** Biochemical testing of *P. thiaminolyticus*. API 50 CHB biochemical results for *P. thiaminolyticus* BKBLC 2156 as previously reported in the case report of Ouyang et al (2010)(*107*), contrasted with our testing of reference type strain NRRL B-4156, and our clinical isolate from patient sample 2033 strain type Mbale. The reference strain NRRL B-4156, and our clinical isolate type strain Mbale, were both identified as *P. thiaminolyticus* with 99.8-99.9% confidence (https://apiweb.biomerieux.com). The clinical strain was tested 3 times, and while there was some variability at 24 h, it consistently identified as P. thiaminolyticus with high confidence at 48 h.

| **Tube #** | **Test** | **Carbohydrate** | **Case report(107)** | **Strain B-4156** | **Clinical Sample 2033 Strain Mbale** |
| --- | --- | --- | --- | --- | --- |
| 0 | Control (NG = no growth) |  | NG | NG | NG |
| 1 | GLY | glycerol | + | + | + |
| 2 | ERY | erythritol | - | - | - |
| 3 | DARA | D-arabinose | - | - | - |
| 4 | LARA | L-arabinose | - | - | - |
| 5 | RIB | D-ribose | + | + | + |
| 6 | DXYL | D-xylose | - | - | - |
| 7 | LXYL | L-xylose | - | - | - |
| 8 | ADO | D-adonitol | - | - | - |
| 9 | MDX | methyl-βD-xylopyranoside | - | - | - |
| 10 | GAL | D-galactose | + | (+) | (+) |
| 11 | GLU | D-glucose | + | + | + |
| 12 | FRU | D-fructose | + | + | + |
| 13 | MNE | D-mannose | + | + | + |
| 14 | SBE | L-sorbose | - | - | - |
| 15 | RHA | L-rhamnose | - | - | - |
| 16 | DUL | dulcitol | - | - | - |
| 17 | INO | inositol | - | - | - |
| 18 | MAN | D-mannitol | - | - | - |
| 19 | SOR | D-sorbitol | - | - | - |
| 20 | MDM | methyl-($\alpha$)D-mannopyranoside | - | - | - |
| 21 | MDG | methyl αD-glucopyranoside | - | (+) | (+) |
| 22 | NAG | N-acetylglucosamine | + | + | + |
| 23 | AMY | amygdalin | + | + | + |
| 24 | ARB | arbutin | + | + | + |
| 25 | ESC | esculin | + | + | + |
| 26 | SAL | salicin | + | + | + |
| 27 | CEL | D-cellobiose | - | (+) | (+) |
| 28 | MAL | D-maltose | + | + | + |
| 29 | LAC | D-lactose (bovine origin) | - | (+) | (+) |
| 30 | MEL | D-melibiose | + | + | + |
| 31 | SAC | D-saccharose | + | + | + |
| 32 | TRE | D-trehalose | + | + | + |
| 33 | INU | inulin | - | - | - |
| 34 | MLZ | D-melezitose | - | (+) | (+) |
| 35 | RAF | D-raffinose | + | + | + |
| 36 | AMD | amidon | + | + | + |
| 37 | GLYG | glycogen | + | + | - |
| 38 | XLT | xylitol | - | - | - |
| 39 | GEN | gentiobiose | + | + | + |
| 40 | TUR | D-turanose | + | + | + |
| 41 | LYX | D-lyxose | - | - | - |
| 42 | TAG | D-tagatose | - | - | - |
| 43 | DFUC | D-fucose | - | - | - |
| 44 | LFUC | L-fucose | - | - | - |
| 45 | DARL | D-arabitol | - | - | - |
| 46 | LARL | L-arabitol | - | - | - |
| 47 | GNT | potassium gluconate | + | (+) | (+) |
| 48 | 2KG | potassium 2-ketogluconate | - | - | - |
| 49 | 5KG | potassium 5-ketogluconate | - | - | - |
| **Likelihood of *P. thiaminolyticus*** |  |  | **57.5%** | **99.9%** | **99.8%** |

NG: No growth - Negative + Positive (+) low / late positive
24 Hours
B-4156
48 Hours

**Supplementary Table S11.** Antibiotic Resistance. Antibiotic Resistance testing of the available antibiotics in the Ugandan population was performed on two clinical isolates of *Paenibacillus* that grew aerobically, using standard E-test sensitivity and interpretations given for ones that were interpretable with Clinical and Laboratory Standards Institute (CLSI) guidelines. The *P. Thiaminolyticus Mbale* isolate, which appeared to represent most of our samples, showed no antibiotic resistance while the *Paenibacillus* spp. was identified as resistant to penicillin with intermediate resistance to clindamycin.

| Antibiotic | *P. thiaminolyticus* |  | *Paenibacillus* spp. |  |
| --- | --- | --- | --- | --- |
|  | MIC (µg/ml) | Interpretation* | MIC(µg/ml) | Interpretation* |
| Vancomycin | 4 | S | 0.125 | S |
| Ciprofloxacin | 0.064 | S | 0.25 | S |
| Penicillin | 0.125 | S | 1.0 | R |
| Clindamycin | 0.5 | S | 2.0 | I |
| Ceftriaxone** | 0.5 |  | 0.5 |  |
| Gentamycin | 2.0 | S | 0.032 | S |
| Tetracycline | 0.25 | S | 0.064 | S |

\*S: Susceptible, R: Resistant, I: Intermediate
\*\* No interpretation by CLSI guidelines

**Supplementary Table S12.** Virulence Testing. Summary of virulence testing. Of the 50 mice inoculated, only those receiving the *Mbale* strain displayed signs of sepsis and were moribund. The infection was only clinically evident at the highest undiluted dose (16/16). Diluted doses were not able to induce signs of infection (0/10).

| **Inoculation Status**<br>**[no. and % in each category]** | **All mice**<br>**n=50** | **Moribund or Died**<br>**n=16** | **Normal**<br>**n=34** |
| --- | --- | --- | --- |
| Mbale strain | 26 (52) | 16 (62) | 10 (38) |
| Undiluted ($\sim 10^9$ CFU/mL) | 16/26 | 15/16* | 0/16 |
| Dilutions (below $10^9$ CFU/mL) | 10/26 | 0/10 | 10/10 |
| Reference strain | 14 (28) | 0 (0) | 14 (100) |
| Normal saline | 10 (20) | 0 (0) | 10 (100) |

\* One mouse infected with the Mbale strain demonstrated signs of illness, but spontaneously recovered after 2 days.

